# Assessing molecular gene by treatment interactions using a population of neural progenitors exposed to valproic acid and lithium

**DOI:** 10.1101/2025.09.12.25335653

**Authors:** Jordan M Valone, Brandon D Le, Nana Matoba, Jessica T Mory, Justin M Wolter, Michael I Love, Jason L Stein

**Affiliations:** Department of Genetics, University of North Carolina at Chapel Hill; Chapel Hill, NC, USA; UNC Neuroscience Center, University of North Carolina at Chapel Hill; Chapel Hill, NC, USA; Carolina Institute for Developmental Disabilities; Carrboro, NC, USA; Department of Biostatistics, University of North Carolina at Chapel Hill; Chapel Hill, NC, USA

**Keywords:** Gene x treatment interactions, neural progenitors, gene regulation, pharmacogenomics, chromatin accessibility, gene expression, quantitative trait loci, valproic acid, lithium

## Abstract

Gene by treatment (GxT) interactions likely contribute to variability in clinical response, but are difficult to identify in population studies. Here, we applied psychiatric and neurological disorder treatments to a genotyped population of human neural progenitors (n=83 donors) and measured molecular responses on chromatin accessibility and gene expression. Gene regulatory responses to valproic acid (VPA), which is also a prenatal risk factor for autism, and lithium were highly enriched in genetic risk for psychiatric disorders, demonstrating the convergence of environmental and genetic factors. Genetic variation impacted molecular response to these drugs at over 1,000 loci, a subset of which modulated the impacts of psychiatric disorder risk variants. Finally, transcriptome-wide association revealed enzymes involved with folate metabolism during VPA exposure impact cognitive ability, a pathway previously shown to alleviate the negative impacts of this exposure. The “GxT in a dish” approach identified a validated treatment pathway, supporting its broad utility.

## Introduction

Variability in clinical response to therapeutic interventions and unintended adverse effects are significant challenges in modern medicine. Pharmacogenomic studies, which examine how an individual’s genetic makeup influences their response to drugs, have had limited success in psychiatry^1,2^, and face inherent difficulties due to confounding variables such as polypharmacy, inconsistent patient adherence to dosing regimens, and the logistical complexity of recruiting large-scale human cohorts. Pharmacogenomic studies have also mostly uncovered genetic effects on the rate of drug metabolism, rather than genetic modifiers of brain cell response to the drug^3^. Understanding the genetic factors that modulate cellular and molecular drug responses remains crucial to advancing personalized medicine and targeted interventions. To move beyond these limitations and enable precision medicine, molecular pharmacogenomic approaches that directly assess the interplay between an individual’s genetic profile and their response to specific treatments or environmental exposures in a controlled cell culture based experiment are required. Here, we apply such an approach to explore the pharmacogenomics of two key treatments for psychiatric and neurological disorders, valproic acid (VPA) and lithium (Li).

VPA is used clinically as an anticonvulsant and mood stabilizer. Prenatal exposure to VPA is also a well-documented environmental cause of teratogenic effects, increasing risks for spina bifida, congenital malformations, autism spectrum disorder (ASD), and intellectual disability^4–6^. Notably, only a subset of exposed individuals are affected. For example, previous studies have found around 10% of individuals prenatally exposed to VPA develop ASD^7^. While the precise determinants of individual susceptibility to these outcomes remain unclear, studies in mice suggest that germline genetic variation likely plays a role in shaping the response to VPA exposure^8^. Despite these risks, VPA may still be prescribed for pregnant mothers when the benefits of treating severe maternal epilepsy or bipolar disorder outweigh the risks of teratogenicity, emphasizing the necessity for better tools to predict and mitigate adverse outcomes^5^. While the precise mechanisms of VPA’s adverse effects are not fully understood, its known actions include histone deacetylase inhibition (HDACi), which can broadly alter gene expression, oxidative stress, and interference with folate uptake through receptor antagonism^9–12^. This latter mechanism is particularly relevant, as disruptions in folate pathways are known to impact neurodevelopment and have been implicated in neural tube defects^13,14^. Molecular pharmacogenomic approaches provide a promising method to investigate the role of an individual’s genetic profile in predicting variability in response.

Similarly, Li is a cornerstone maintenance treatment for bipolar disorder, though has variable patient outcomes^2,15–19^. This variability is likely partially driven by genetic background shared with bipolar disorder risk, as indicated by family studies and emerging polygenic evidence^2,17,20^. Despite pharmacogenomic studies in patient populations, the specific genetic underpinnings of Li responsiveness remain largely unknown, in part due to the previously mentioned challenges of sufficient power and limited control of exposures. A hypothesis for Li’s therapeutic effects centers on its capacity to stimulate neurogenesis^21,22^, which has previously been studied in human neural progenitors cell (hNPC) and induced pluripotent stem cell (iPSC)-derived neural cultures from Li responders and non-responders^23,24^. The ongoing challenge in identifying robust genetic markers for both VPA and Li response underscores the need for controlled experimental models.

To address these challenges, we employed a “GxT in a dish” approach utilizing a population of genotyped human neural progenitor cells with inherent genetic variation. As a living and homogeneous cellular model of brain development, hNPCs offer a powerful platform to assess gene regulatory responses to well-controlled exposures, circumventing many of the confounding variables inherent in human cohort pharmacogenomic studies^25–28^. This model allows for the direct observation of how genetic variation influences molecular responses to clinically relevant exposures, thereby revealing context-dependent genetic effects that would otherwise remain undetected^29^. By exposing hNPCs from 83 genotyped donors to VPA and Li, we aimed to uncover novel regulatory functions of genetic variants and characterize GxT interactions.

Our study demonstrates that both VPA and Li induce widespread and distinct epigenomic and transcriptomic changes in hNPCs, with VPA notably shifting their state from proliferation towards differentiation, while Li promotes proliferation. We identified hundreds of genetic variants whose impact on gene regulation was evident only under VPA or Li exposure, which were more often distal from promoters and affected loss of function intolerant genes. By integrating these genetic findings from each drug treatment with existing genome-wide association study (GWAS) data, we find that VPA can accentuate molecular effects underlying neuropsychiatric disorder risk and cognitive deficits. Using a transcriptome-wide association study (TWAS), we found that the effects of VPA on cognitive ability are mediated by a convergence of environmental and genetic factors within the folate metabolism pathway. This finding provides a specific molecular mechanism consistent with animal models and clinical observations and suggests a target for therapeutic intervention^14,30^. This “GxT in a dish” framework thus provides a robust strategy for functionally characterizing the complex interplay between genetic predisposition and environmental factors, demonstrating a strategy that could expand the possibilities of personalized medicine.

## Results

### Measuring alterations in gene regulation in hNPCs due to VPA and Li

We investigated gene regulatory signals in neural progenitors from a population of genotyped, multi-ancestry hNPC donors after exposure to clinically useful drugs. After a two week expansion period, we exposed hNPCs from 83 unique donors to vehicle, 1mM VPA, or 1.5mM Li treatment for 48h. VPA and Li exposure concentrations were chosen to approximate clinically relevant levels^19,31–34^. The 48-hour time point was chosen based on its robust impacts on molecular and cellular responses in prior research, allowing the study of acute effects^24,35,36^. Cells were subsequently collected for epigenomic and transcriptomic profiling via ATAC-seq and total RNA-seq (Fig. 1a). Technical reproducibility was assessed through the inclusion of 2-5 replicates for 6 randomly selected donors, demonstrating high consistency (Supplementary Fig. 1). After filtering samples based on quality control (Methods), we detected expression of 15,438 protein-coding genes and 7,241 long non-coding RNAs (lncRNAs) from 232 RNA-seq samples (n_veh_ = 76, n_VPA_ = 78, n_Li_ = 78) and accessible chromatin at 172,354 peaks from 232 ATAC-seq samples (n_veh_ = 76, n_VPA_ = 75, n_Li_ = 81).

**Fig. 1.**
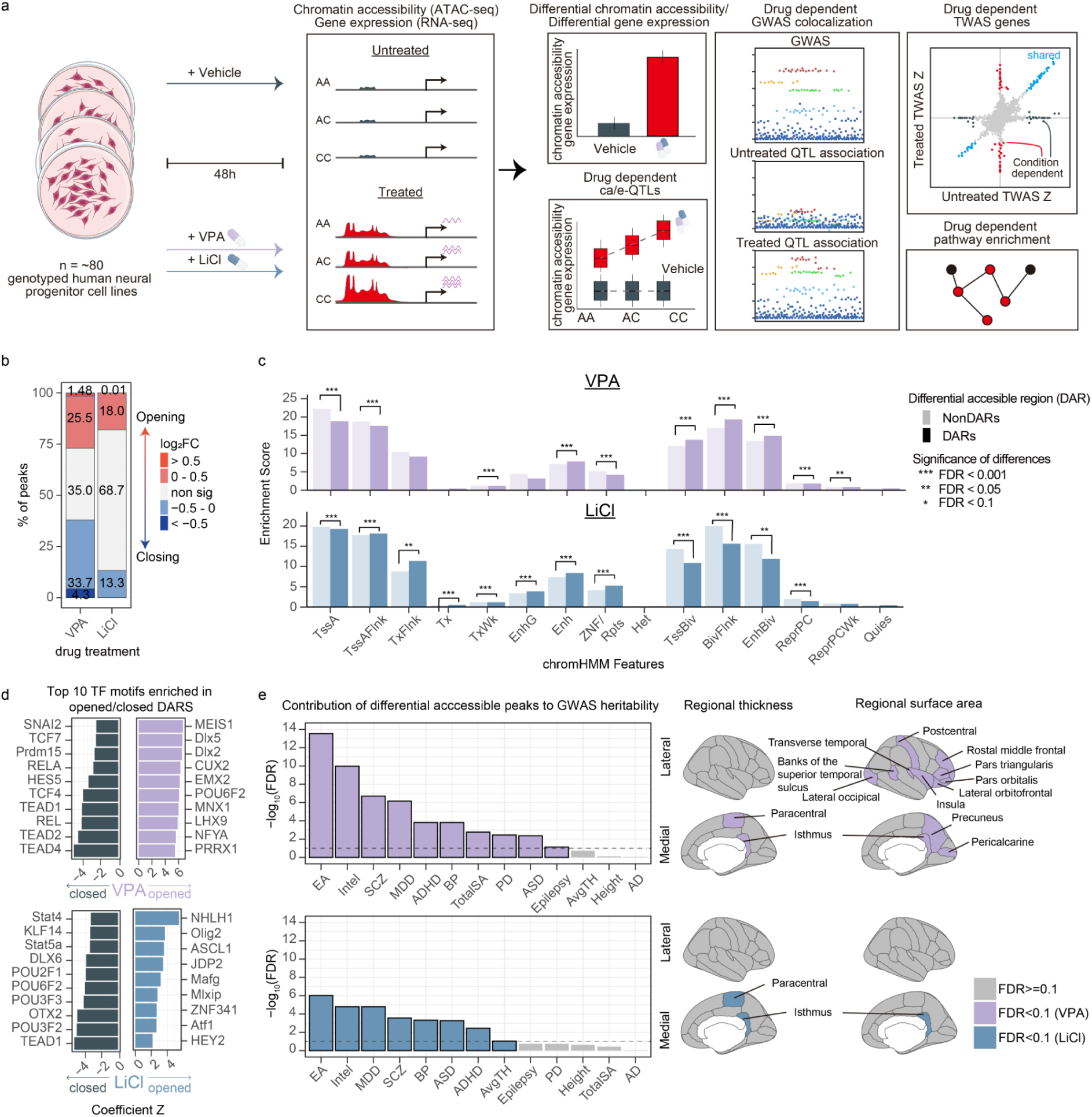
Chromatin accessibility changes due to drug exposure. (a) Experimental design cartoon. (b) Inventory of differentially accessible regions (DARs) for VPA or Li relative to vehicle annotated by effect direction and magnitude. (c) Enrichment of chromHMM^40^ features (c) within VPA (top) or Li (bottom) DARs and non-DARs. Asterisks indicate a significant difference between DARs and non-DARs after multiple testing correction. (* = FDR < 0.1, ** = FDR < 0.05, *** = FDR < 0.001). (d) Top 10 TFBS motifs significantly enriched within closed (left) or opened DARs. (e) Contribution of DARs to heritability of neuropsychiatric risk, cognition, brain structure, and other complex traits.

### Changes in chromatin accessibility induced by VPA or Li

We performed differential chromatin accessibility and gene expression analyses that showed VPA or Li treatment induced broad epigenomic and transcriptomic changes relative to vehicle treatment. We defined significantly differentially accessible chromatin regions (DARs) as those with a false discovery rate (FDR-adjusted *P*) below 0.1 (Supplementary Table 1). VPA treatment had a strong effect on the epigenome, inducing 111,978 DARs, which represent 65% of all chromatin peaks. While VPA functions as a histone deacetylase (HDAC) inhibitor^10^, typically associated with chromatin opening, our findings revealed that VPA DARs were more often closed than opened (58% closed DARs, Binomial test *P* < 9.9 x 10^-324^) (Fig. 1b). This observation aligns with prior studies that show the impact of VPA on chromatin organization is highly cell-type specific^36,37^. In contrast, Li treatment induced smaller effect sizes and fewer DARs (30,613 DARs), representing 18% of all chromatin peaks genome-wide. Li DARs tended to open rather than close (54% opened DARs, Binomial test *P* < 1.7 x 10^-41^) (Fig. 1b), which is consistent with previous findings(^38,39^). We detected distinct epigenomic alterations induced by VPA and Li, potentially challenging conventional views of their chromatin remodeling effects.

To confirm that our DARs represent putatively functional regulatory regions of the genome, we tested hNPC chromatin profiles for enrichment with chromatin state annotations provided by chromHMM^40^(Fig. 1c; Supplementary Tables 2 and 3). Across both DAR and non-DAR peak sets we found enrichments for features related to transcription start site (TSS) and enhancers, but not heterochromatin, quiescent or transcribed regions, consistent with previous findings for open chromatin regions^41^. Interestingly, we found VPA DARs showed a significantly greater enrichment (FDR-adjusted *P* < 0.1) for bivalent markers when compared to non-DAR elements. Bivalent sites are defined by both activating and repressive markers and have been shown to regulate genes related to developmental processes^42^. These VPA-induced shifts at sites which are usually bivalent in developing brain tissue may impact fate decisions in these cells.

DARs alter gene regulation by regulating access to transcription factor (TF) binding sites within open or closed regulatory regions. To implicate specific TFs and their downstream gene regulatory networks as molecular mediators of VPA or Li response in hNPCs, we analyzed the top 2000 most opened or closed DARs for enrichment of 841 unique TF binding site (TFBS) motifs from the JASPAR database^43^ (Fig. 1d; Supplementary Table 4). Within VPA DARs, TFBS motifs associated with proliferative cellular processes, such as those related to the WNT pathway, including the TCF7 motif, were significantly enriched in closed DARs. Conversely, motifs favoring differentiation and neurogenesis, like Cux2 and Dlx5, were significantly enriched in opening DARs^44,45^. This finding is consistent with VPA inducing a developmental shift in human neural progenitors, promoting neuronal differentiation while suppressing their proliferative capacity. Pathway enrichment analysis of TFs associated with VPA-responsive DARs further supported these trends (Supplementary Fig. 2; Supplementary Table 5). Terms such as “Hippo signaling pathway”, associated with TEAD, and “Wnt signaling pathway”, associated with TCF, were significantly enriched among TFs associated with VPA-closing DARs, further supporting the inhibition of proliferation pathways^46–48^. Similarly, “neuron differentiation” was significantly enriched among TFs associated with VPA-opening DARs. Li-responsive DARs similarly implicated TFBS motifs also related to differentiation but in an opposing pattern. We observed a significant enrichment in opening DARs for transcription factors related to proliferative fate decisions, such as HEY2 and ASCL1^49,50^. The pathway enrichment analysis for Li-responsive DARs also corroborated these findings. Specifically, GO terms like “axonogenesis” and “neuron projection morphogenesis” were significantly enriched among TFs associated with Li-closing DARs, suggesting that Li treatment suppresses neuronal fate decisions. Overall, both the individual TF motif analysis and the comprehensive pathway enrichment results consistently highlighted opposing effects of VPA and Li on neural progenitor fate decisions.

### DARs linked to brain trait heritability

Prenatal VPA exposure is a known risk factor for ASD, cognitive impairments, and microcephaly motivating investigation into whether an environmental risk factor and genetic risk factors have convergent effects. To evaluate whether DARs showed enrichment of SNPs conveying heritability for cognitive traits, neuropsychiatric disorder risk, or variability in cortical structures assayed through GWAS, we applied stratified linkage disequilibrium score regression^51,52^ (Fig. 1e; Supplementary Table 6). Intelligence, educational attainment (a proxy for cognitive ability), and risk for neuropsychiatric disorders showed the highest heritability enrichments in VPA DARs. We also found that both VPA DARs showed enrichment for ASD risk. In contrast, VPA DARs were not enriched for height, a strongly polygenic non-brain specific trait, or for neurodegenerative disorders like Alzheimer’s disease risk, though we detected modest enrichment for Parkinson’s disease risk within VPA DARs. These findings suggest a convergence of genetic and environmental factors, where genetic vulnerabilities for cognitive traits are enriched in regions perturbed by environmental influences.

Additionally, we observed partitioned heritability enrichment for diseases where the drug is a primary treatment. Specifically, Li DARs showed significant heritability enrichment for bipolar disorder. This finding is particularly relevant given previous associations of genetic variability influencing inter-individual differences in response to Li induced proliferation colocalizing with GWAS signal for bipolar disorder, suggesting these Li DARs may encompass genetic factors influencing both disease susceptibility and related treatment outcomes^24^. Similarly, VPA DARs showed an enrichment for bipolar disorder and showed slight enrichment for epilepsy, a condition for which VPA is also used as a therapeutic. This suggests that drug perturbations impact chromatin regions relevant to disease etiology.

Building on the established neurodevelopmental consequences of VPA, such as microcephaly, and the capacity of Li to promote neural progenitor proliferation, we next focused our analysis on cortical brain structures. We found significant enrichment of heritability of total cortical surface area within VPA DARs, consistent with prenatal VPA exposure impacting brain size (Fig. 1e)^53,54^. We also found enrichment of either cortical thickness or surface area for 13 cortical regions, where, interestingly, surface area enrichments were largely in the frontal cortex, suggesting regional susceptibility of VPA exposures in areas important for cognition and language. We also found enrichment for 2 cortical regions within Li DARs (Fig. 1e). Both Li and VPA DARs displayed enrichment for SNPs affecting thickness of the paracentral cortex, a region also associated with changes in thickness and surface area in bipolar patients taking Li or antiepileptic medications, where Li-treated bipolar disorder patients showed increased thickness and surface area, and antiepileptic drug-treated bipolar disorder patients showed decreased thickness^55^. Collectively, these results highlight a convergence of environmentally induced changes in gene regulation with common genetics underlying risk for psychiatric disorders, inter-individual differences in cognitive ability, and cortical structure.

### Changes in gene expression and proliferation induced by VPA or Li

Given that VPA and Li induce widespread changes in chromatin accessibility, a key mechanism regulating gene expression, we next investigated differential expression induced by either treatment. We identified differentially expressed genes (DEGs), defined as those with significantly different expression under drug exposure conditions relative to vehicle conditions (FDR-adjusted *P* < 0.1), detecting over 15,000 and 10,000 genes in response to VPA or Li treatments, respectively (Supplementary Table 7). Notably, the magnitude of gene expression changes was considerably larger following VPA treatment compared to Li treatment, a finding consistent with the greater effect sizes observed in chromatin accessibility after VPA exposure. As VPA is a known inhibitor of histone deacetylase activity, we examined differential expression of HDACs themselves. We found that most HDAC genes, including HDAC class I and IIa that are direct targets of VPA, were upregulated due to VPA exposure, possibly as a compensatory mechanism ^10,56^(Supplementary Fig. 3). Further analysis of these DEGs revealed distinct patterns related to cellular proliferation. Specifically, genes associated with cellular proliferation, such as cell cycle and DNA replication pathways, were upregulated by Li and downregulated by VPA (Fig. 2 a,b). For example, *CDK1*, a master regulator of the G2 to M phase transition, and the transcription factor *E2F1*, which drives S-phase progression, were both found to be upregulated by Li and downregulated by VPA^57,58^. Similarly, core components of the DNA replication machinery, such as the MCM family of genes were also found to exhibit this pattern of expression^59^. *GNL3* was also identified as a DEG that was upregulated by Li and downregulated by VPA, consistent with its previously reported role in driving lithium-induced proliferation^24^. These observed changes in gene expression are consistent with chromatin accessibility findings described above.

**Fig. 2.**
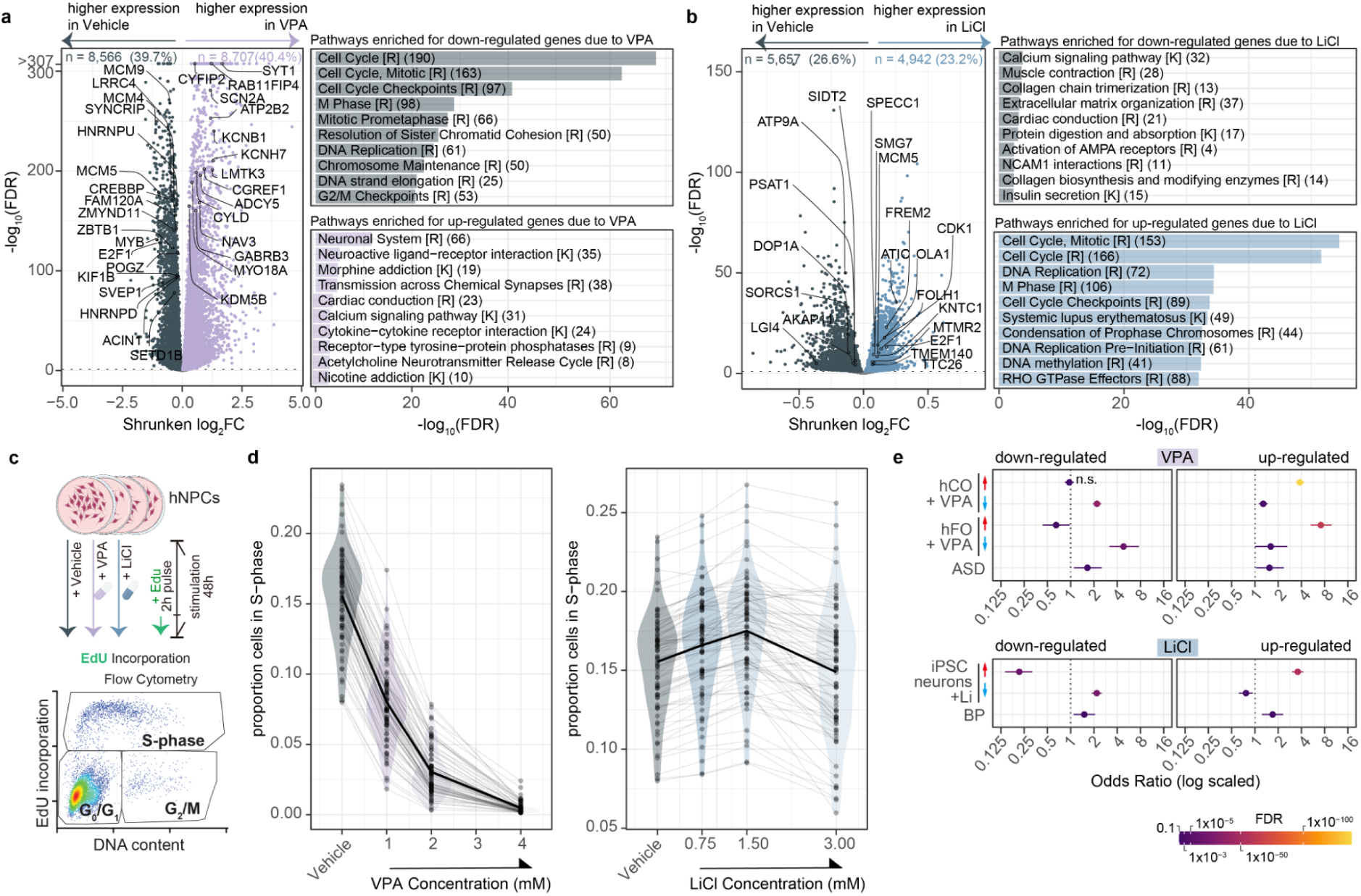
Differential gene expression and proliferation effects due to VPA and Li. Volcano plots of differentially expressed genes (DEGs) induced by VPA relative to vehicle (a, left) and Li relative to vehicle (b, left). Positive and negative log_2_FC values reflect higher or lower gene expression in the drug treated condition relative to vehicle, respectively. Pathway enrichment within VPA (a, right) or Li DEGs (b, right). Numbers in parentheses indicate the intersection size, i.e., the number of DEGs overlapping with each pathway. K = KEGG, R = Reactome. (c) Proliferation assay experimental design. (d) Proliferation assay results for VPA (left) and Li (right, previously published in ^24^) displaying the proportion of cells in S-phase across each donor, and the mean across all donors for a given concentration in bold. (e, top) Fisher’s exact test for overlap between up-regulated (up arrow) or down-regulated (down arrow) VPA-DEGs and DEGs from VPA-treated human brain organoids^35,60^ or ASD-associated genes^62^. (e, bottom) Fisher’s exact test for overlap between Li-DEGs and DEGs from Li-treated human iPSC neurons^61^ or BIP-associated genes^63^. In (e), lines represent the 95% confidence interval associated with the log-scaled odds ratio estimates. hCO = human cortical organoid, hFO = human forebrain organoid, “n.s.” indicates a non-statistically significant result.

To validate the effects on proliferation, we employed an 5-ethynyl-2’-deoxyuridine (EdU) assay, which incorporates a nucleoside analog to label newly synthesized DNA during S-phase of the cell cycle, enabling the quantification of proliferating cells via flow cytometry when combined with a DNA content dye (Fig. 2c). We utilized this assay in 80 donors to quantify response to drug and inter-individual variability in response (Fig. 2d). As shown in our previous work^24^, Li exposure has a dose-dependent effect on proliferation with clinically relevant concentrations^19,31^ (0.75-1.5 mM) increasing proliferation while higher concentrations decrease proliferation. In contrast, VPA exposure leads to consistent reductions in proliferation across all donors at clinically relevant concentrations^32–34^ (1 mM), and to an almost complete shutdown of proliferation at the highest concentrations tested. These findings strongly support the gene expression and chromatin accessibility findings.

To assess the reproducibility of drug-induced DEGs, we examined the overlap between our identified VPA-DEGs and Li-DEGs and previously reported differentially expressed genes from VPA-treated human brain organoids^35,60^ and Li-treated human iPSC neurons^61^, respectively, using Fisher’s exact tests (Fig. 2e). This analysis revealed significant and robust overlap for the majority of comparisons, suggesting that the observed changes in gene expression represent reproducible biological responses across studies. We also tested the overlap between our VPA-DEGs and known ASD-associated genes^62^, and between Li-DEGs and BIP-associated genes from whole exome sequencing studies^63^. This revealed significant overlaps, again illustrating a convergence of environmental exposure and genetic factors, where exposure-induced gene expression changes align with genes implicated in neuropsychiatric disorders, consistent with our findings for chromatin accessibility.

### Regulatory elements with drug specific effects on gene expression

To infer regulatory elements impacting gene expression within each drug, we integrated our chromatin accessibility and gene expression data. We quantified this association by correlating chromatin accessibility peaks with the expression of genes located within 1 Mb of their transcription start sites (TSS). This analysis revealed that over 11% of peaks correlated with nearby genes, and nearly 50% of genes correlated with nearby peaks across both exposed and vehicle conditions (FDR-adjusted *P* < 0.1, median peak–TSS distance ∼100 kb; Supplementary Fig. 4a-c, Supplementary Fig. 4a-c, Supplementary Table 8). A positive correlation was observed in over 75% of gene-peak pairs, which is consistent with the principle that increased chromatin accessibility generally leads to higher target gene expression^28,29^. We found that most significantly correlated (FDR-adjusted *P* < 0.1) gene-peak pairs were condition-specific: 5,174 were unique to the vehicle condition, 5,540 to the VPA-exposed condition, and 5,885 to the Li-exposed condition. In contrast, 3,975 gene-peak pairs exhibited significant correlation across all conditions (Supplementary Fig. 4d). The VPA DEG *NRXN3* is a notable example of drug-dependent changes in gene regulation, where VPA induced increased gene expression and 13 additional significant peak-gene correlations emerged exclusively after VPA exposure (Supplementary Fig. 4e). *NRXN3* plays a role in neuronal differentiation, which is consistent with our findings on the impacts of VPA exposure, and is linked to ASD, aligning with a known risk of VPA exposure^64,65^. Pathway enrichment analysis showed that VPA-specific gene-peak pairs were significantly enriched for neurexins and neuroligins, framing the regulatory shifts at *NRXN3* as part of a broader process of inducing neuronal differentiation and synapse formation(Supplementary Fig. 4f). These findings suggest that distinct sets of regulatory elements are largely active after exposure to drugs to control gene expression.

### Genetic effects on gene regulation altered by VPA or Li

Having found gene regulatory changes caused by each drug, we next aimed to find SNPs associated with gene regulation within each drug exposure to understand inter-individual variability in response. We measured the effects of common genetic variants within each exposure by mapping chromatin accessibility and gene expression quantitative trait loci (caQTLs and eQTLs, respectively). We detected over 24,000 distinct caQTL signals (caSNP-caPeak pairs) in each exposure regulating 20,596 unique caPeaks (FDR-adjusted *P* < 0.1; number of caQTL pairs: 26,438 vehicle; 24,470 VPA; 28,706 Li; Fig. 3a,b; Supplementary Table 9). We also found over 1,900 eQTLs (eSNP–eGene pairs) in each exposure regulating 3,037 unique eGenes (FDR-adjusted *P* < 0.1; number of eQTL pairs: 1,936 vehicle; 1,979 VPA; 2,022 Li; Fig. 3c,d; Supplementary Table 10).

**Fig. 3.**
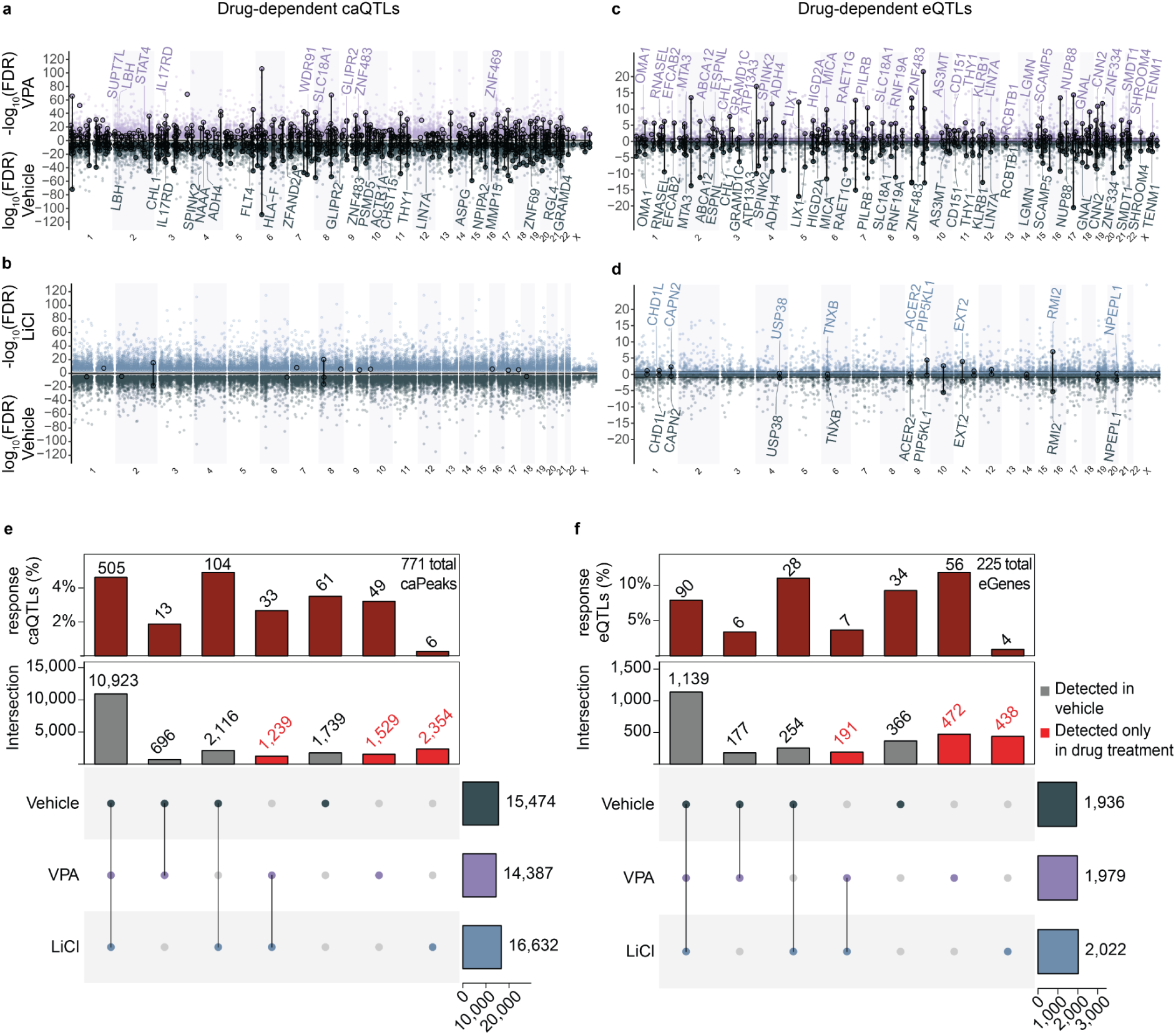
Genetic Variation Impacts Molecular Response to Drug Exposure. Miami plots showing significant caQTLs (a, b) or eQTLs (c, d) identified under VPA (purple; a, c), Li (blue; b, d), or vehicle (gray) conditions. Circled variants highlight significant genotype-by-condition interaction effects (r-QTLs). Labels depict a subset of protein-coding eGenes which overlap drug-dependent ca/eQTLs. UpsetR plots illustrate the sharing of caPeaks (e) and eGenes across drug exposures (f). Red-labeled columns denote those caPeaks or eGenes identified solely in drug treated conditions. The top panel shows the percentage of drug-dependent caPeaks (e) or eGenes (f) in each category found to be r-QTLs.

While most QTLs were identified consistently regardless of drug treatment, the functional impact of some variants was evident only under drug exposures. caQTLs identified exclusively in VPA or Li exposures altered the accessibility of 1,529 and 2,354 chromatin peaks, respectively (Fig. 3e). Similarly, eQTLs found only after exposure modulated the expression of 472 and 438 distinct genes in VPA and Li conditions, respectively, demonstrating that genetic variation has detectable impacts on a substantial number of genes specifically when exposed to these drugs (Fig. 3f).

We also performed genotype-by-condition interaction tests to further characterize the drug-dependent function of genetic variation (response-QTLs, r-QTLs). This approach provides a statistically supported assessment of how treatments alter genetic effects. We detected 779 and 15 r-caQTLs in VPA, and 214 and 15 r-eQTLs in Li (Supplementary Tables 11 and 12). Because interaction tests require increased power relative to standard genetic associations, these counts reflect the most robust interaction signals, but do not rule out the existence of other drug-dependent effects that could be discovered with higher sample sizes^66^. r-QTLs altered both the effect of QTLs detected in vehicle and QTLs only detected during treatment, implying both modulated and novel drug-dependent regulatory mechanisms (Fig. 3e,f). Across both caQTL and eQTL datasets we found strong correlations between vehicle and drug condition effect sizes, in both magnitude and directionality, at common sites. However, we consistently observed lower correlations in effect sizes at r-QTLs, as expected when for interaction effects (Supplementary Fig. 5). This further illustrates both drug-dependent modulation of regulation present under vehicle conditions as well as distinct regulatory mechanisms after drug exposure. One VPA r-caQTL was found to co-localize with an eQTL for *STAT4* (Supplementary Fig. 6 and Supplementary Table 13). This example is particularly interesting as this gene is part of the JAK-STAT signaling pathway which has previously been reported to be affected by VPA exposure^48,67^. Our results provide evidence of novel genetically regulated elements and genes under exposure to clinically relevant conditions while also revealing genetic effects on gene regulation that vary with specific drugs. Both would go undetected in untreated cells highlighting the importance of this “GxT in a dish” approach.

### Characteristics of Condition Dependent Regulatory QTLs

We next sought to examine genomic characteristics of our condition-dependent signals. To do this, we aggregated data on context-dependent WNT regulatory signals from our previous study^29^ with the data generated here. This data included stimulation of hNPCs by CHIR, a small molecule that activates WNT signaling by inhibiting GSK3β, and WNT3A, a recombinant protein that directly stimulates the WNT pathway as a ligand. First we determined whether our drug defined r-QTLs were also identified in other exposure contexts (VPA, Li, CHIR, and WNT3A). For both r-caQTL and r-eQTL, the majority of our signals were found to be unique to a single context, largely driven by VPA (Fig. 4a,b; Supplementary Table 14). Less than 17% and 8% of response caQTL and eQTL, respectively, were found in more than one context. No responsive chromatin sites were common across all of our contexts, and only one was found across all four contexts in response eQTLs, demonstrating that response QTLs exhibit a high degree of context-specificity.

**Fig. 4.**
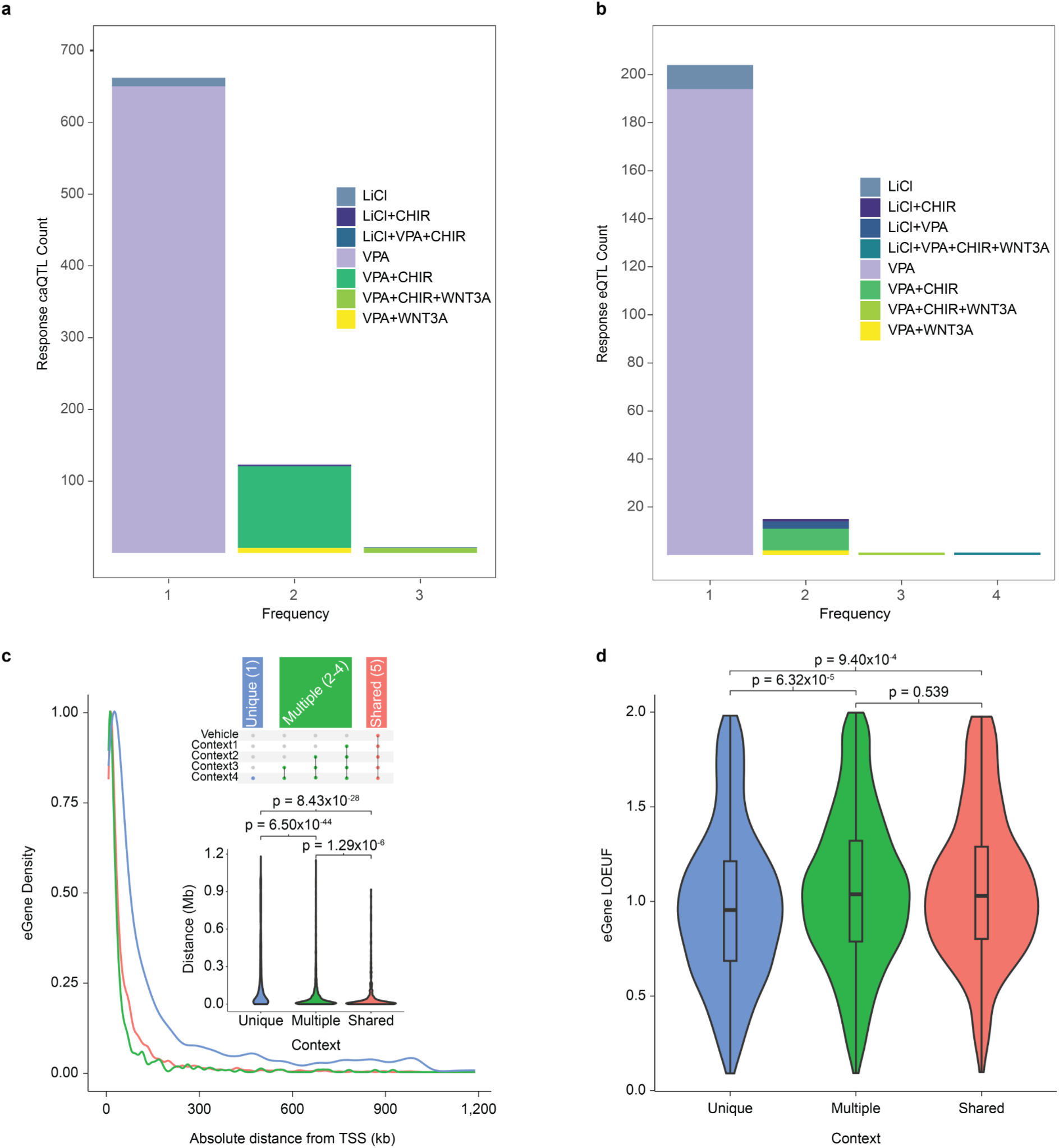
Characteristics of Condition-Dependent QTLs. Bar plots depicting frequency of r-QTL sharing across contexts for r-caQTL (a) and r-eQTL (b). Colored segments represent various context combinations (see legend). (c) Distributions representing the absolute genomic distances between different eQTL groupings (unique, multiple, and shared) and the TSS of their target gene. Inset violin plots display a summary of these distances, accompanied by FDR-adjusted p values. (d) Violin plots showing LOUEF distributions across the same eQTL groups along with FDR-adjusted *P* values. *n_Unique_* = 1,506, *n_Multiple_* = 2,577, *n_Shared_* = 328. Two-sided *P* values were obtained by Wilcoxon rank-sum test (c, d).

To further explore genomic features associated with context specificity, we grouped the lead eQTL variants across all datasets into three categories: unique, multiple, and shared based on the number of contexts in which they were detected (Methods; Supplementary Table 15). All group pairings were found to have a significantly different distance to the TSS of regulated eGenes (Fig. 4c). In particular, eQTLs observed in fewer contexts (Unique > Multiple > Shared) are more distal to the TSS. Our results align with the hypothesis that regulatory elements that function in a context-dependent manner are situated at a greater distance from the genes they regulate compared to shared regulatory elements^68–71^.

We also compared loss-of-function observed/expected upper bound fraction (LOEUF) scores from the Genome Aggregation Database (gnomAD)^72^ between genes with unique, multiple, or shared eQTLs across conditions (Fig. 4d). The purpose of this metric is to demonstrate a gene’s sensitivity to loss-of-function (LoF) variation. A lower score represents a depletion of observed LoF variation relative to expected mutation rate and suggests a higher selective constraint of that gene. Genes affected by variants in only one context were more likely to be loss of function intolerant with lower LOEUF scores relative to the other two groups, suggesting these genes are under stronger negative selection. Previous work has shown strong correlation between low LOEUF scores and disease associated genes^72^. Our results suggest that the regulatory effects found exclusively in a single context are associated with genes under stronger selective pressure and are more likely to be linked to disease.

### Drug-dependent Regulatory Variants Linked to Brain Phenotypes

While partitioned heritability analysis demonstrated enrichment of neuropsychiatric disorder risk and brain structure GWAS loci within drug-induced DARs (Fig. 1), these enrichments do not specify the genes, peaks, or variants contributing to differences in responses to drugs. To pinpoint specific variants and gene regulatory mechanisms where genetic variation alters response to drugs leading to differences in brain traits, we conducted an LD-based overlap analysis of caQTLs and eQTLs with GWAS loci (Supplementary Fig. 7; Supplementary Table 16). This analysis revealed 1,305 caPeak regulatory elements and 181 eGenes under vehicle conditions overlapping at least one brain trait associated locus (Fig. 5a). After considering drug-treated conditions the number of unique brain trait associated peaks and genes increased by 39% and 61% respectively, suggesting drug exposures may accentuate or mask gene regulatory effects at brain-trait associated variants (Fig. 5b). Overall, the detection of 4,432 caPeak–GWAS pairs and 1,152 eGene–GWAS pairs occurred exclusively in drug-treated conditions (Fig. 5c and Supplementary Tables 17 and 18). This overlap supports the hypothesis that genetic variation impacts variability in brain traits by altering molecular response to drugs.

**Fig. 5.**
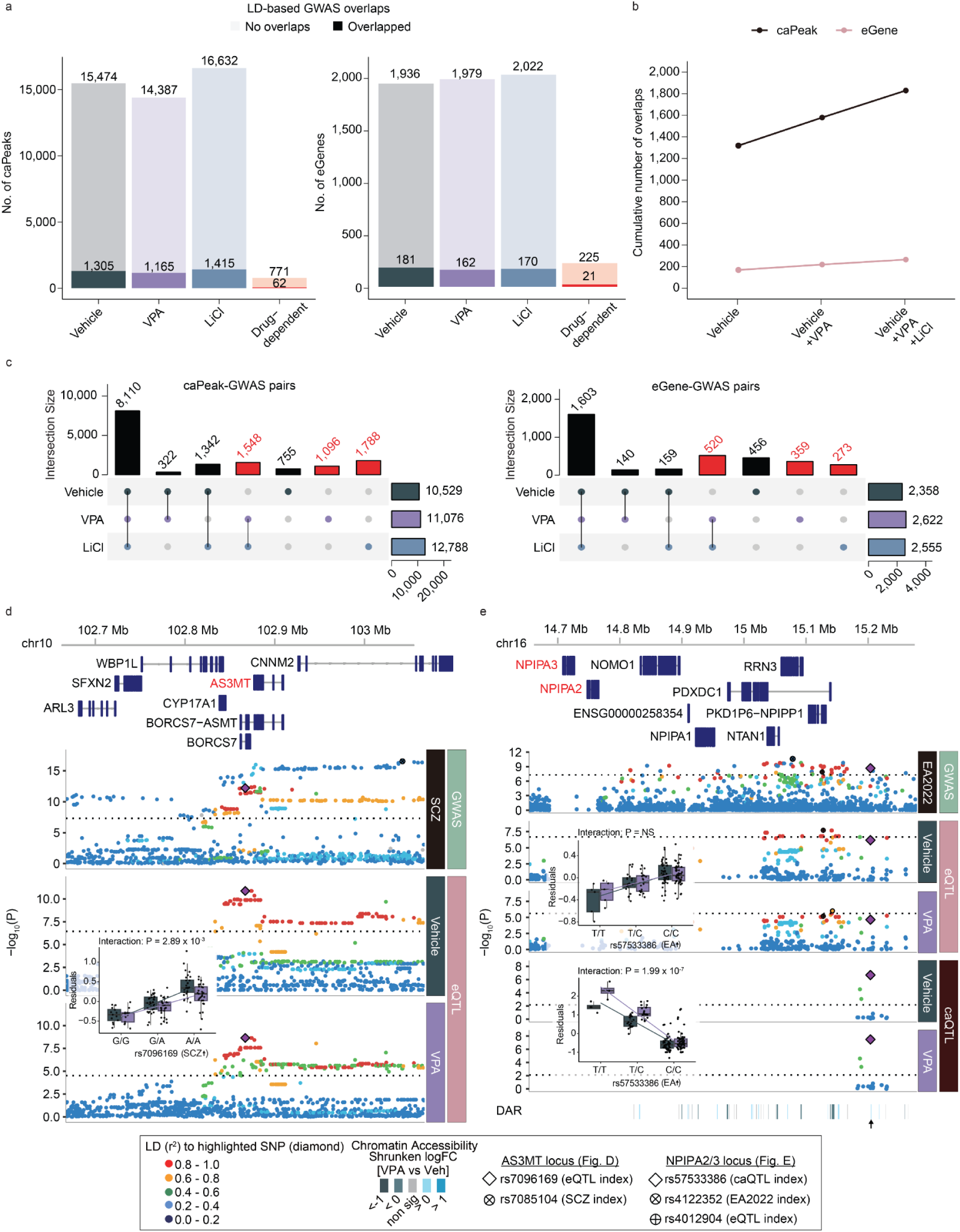
Genetic Variation Altering Molecular Response to Drugs at Brain Trait Associated Loci. (a) Overlap of caPeaks (left) or eGenes (right) with brain-related GWAS loci defined by moderate LD (*r^2^* > 0.6 in either 1KG EUR population or our study) across vehicle, VPA, and Li conditions, as well as for interaction significant drug-dependent caPeaks/eGenes (see Supp Table 16 for brain-trait GWAS). (b) Increasing number of brain traits overlapping caPeaks or eGenes when considering drug-treated conditions. (c) Overlap of caPeaks (left) and eGenes (right) with brain-related GWAS traits in each condition. Overlaps detected exclusively in drug conditions indicated by red columns. (d) Regional association plot showing the co-localization of schizophrenia GWAS^74^ (top panel) with an AS3MT-modulating VPA-responsive eQTL (rs7096169). From top to bottom, the panels display genomic coordinates and gene models, *P* values for brain-related GWAS, and *P* values for QTLs found in this study. (e) Regional association plot illustrating the co-localization of educational attainment GWAS^79^ (top panel) with an eQTL modulating *NPIPA2* and *NPIPA3* expression (*NPIPA3* represented in figure) and a VPA-responsive caQTL (rs57533386; chr16:15206521-15207310), arranged as in (d) *P* values for caQTLs were calculated using a likelihood-ratio test in RASQUAL. *P* values for eQTLs were determined via a linear mixed model with a two-sided test. GWAS *P* values were obtained from the original GWAS summary statistics. QTL boxplots indicate FDR-adjusted interaction *P* values (d,e) (Methods). NS, not significant; SCZ, schizophrenia; EA, educational attainment.

For interaction-significant drug effects (r-QTLs), we found 62 caPeaks and 21 eGenes that had LD overlap with at least one brain trait-associated GWAS locus. We further highlight three of these sites which also met eCAVIAR colocalization criteria (CLPP > 0.01)^73^ (Supplementary Tables 19 and 20). First, a VPA r-eQTL for *AS3MT* (rs7096169-A; Fig. 5d) co-localized with a schizophrenia GWAS signal (rs7085104-A)^74^. We detected a responsive suppression of *AS3MT* expression after VPA treatment relative to vehicle at the r-QTL index variant (rs7096169-A), which was associated with increased risk for schizophrenia. A previous study linked the schizophrenia risk allele to lower risk for epilepsy and higher risk of adverse drug reactions in children treated with VPA, supporting a potential human pharmacogenomic impact though discovered in low sample sizes^75^. AS3MT is a methyltransferase that detoxifies arsenic using S-Adenosylmethionine (SAM) as the methyl donor, which may be affected by VPA-induced disruption of folate metabolism^76^. Another example is a VPA r-caQTL (rs57533386; Fig. 5e) co-localized with an eQTL site for both *NPIPA2* and *NPIPA3*, members of a gene family under strong positive selection in humans and found in the 16p13.11 region, where microdeletions are associated with autism and intellectual disability ^77,78^. This responsive chromatin site also co-localized with an educational attainment GWAS locus^79^. We found a decrease in chromatin accessibility and increase in *NPIPA2/3* expression at rs57533386-C, indicating increased accessibility of this regulatory element is associated with decreased expression. We found that increased chromatin accessibility in the VPA exposure condition accentuated the impacts of the educational attainment decreasing allele, but did not significantly alter gene expression.

Additionally, we highlight a VPA r-caQTL (rs6718978) which co-localized with an eQTL for the *SUPT7L* gene as well as a GWAS locus for cortical thickness (rs6738528; Supplementary Fig. 8). While there was no significant effect of the variant on *SUPT7L* gene expression without drug exposure, a significant increase in expression and a concurrent modulation of chromatin accessibility at a nearby regulatory element were observed after VPA exposure. Specifically, the rs6718978-G allele associated with increased cortical thickness, decreased chromatin accessibility, a relationship that was significantly accentuated by VPA exposure. Notably, *SUPT7L* encodes a protein subunit of the STAGA complex, a known histone acetyltransferase^80^, further supporting a regulatory shift related to histone acetylation. Overall, these results indicate that genetic variation at disease and pharmacogenomic relevant loci alter the response to clinically relevant drugs.

### TWAS highlights drug-dependent gene-trait associations

To further investigate the impact of genetic variation altering response to drugs, we implemented a TWAS approach^81^ which gains power relative to colocalizations by constructing gene expression models based on QTLs detected in each drug treatment across all proximal variants and integrating with GWAS. Brain trait GWASs were selected based on previously reported relevance for VPA (ASD, Epilepsy, Education attainment, Intelligence, cortical surface area and thickness) or Li (bipolar disorder) (Fig. 6, Supplementary Fig. 9a-b, Supplementary Table 21). Consistent with the colocalization analysis (Supplementary Fig. 8), *SUPT7L* was captured in TWAS for cortical thickness as well as educational attainment and intelligence under VPA treatment, strengthening the evidence for its involvement. We also found 105 genes significantly associated with bipolar disorder GWAS under Li treatment including the previously reported genes such as *FADS1*, *TRANK1*, *BDNF*^82–84^ (Supplementary Fig. 9b,c).

**Fig. 6.**
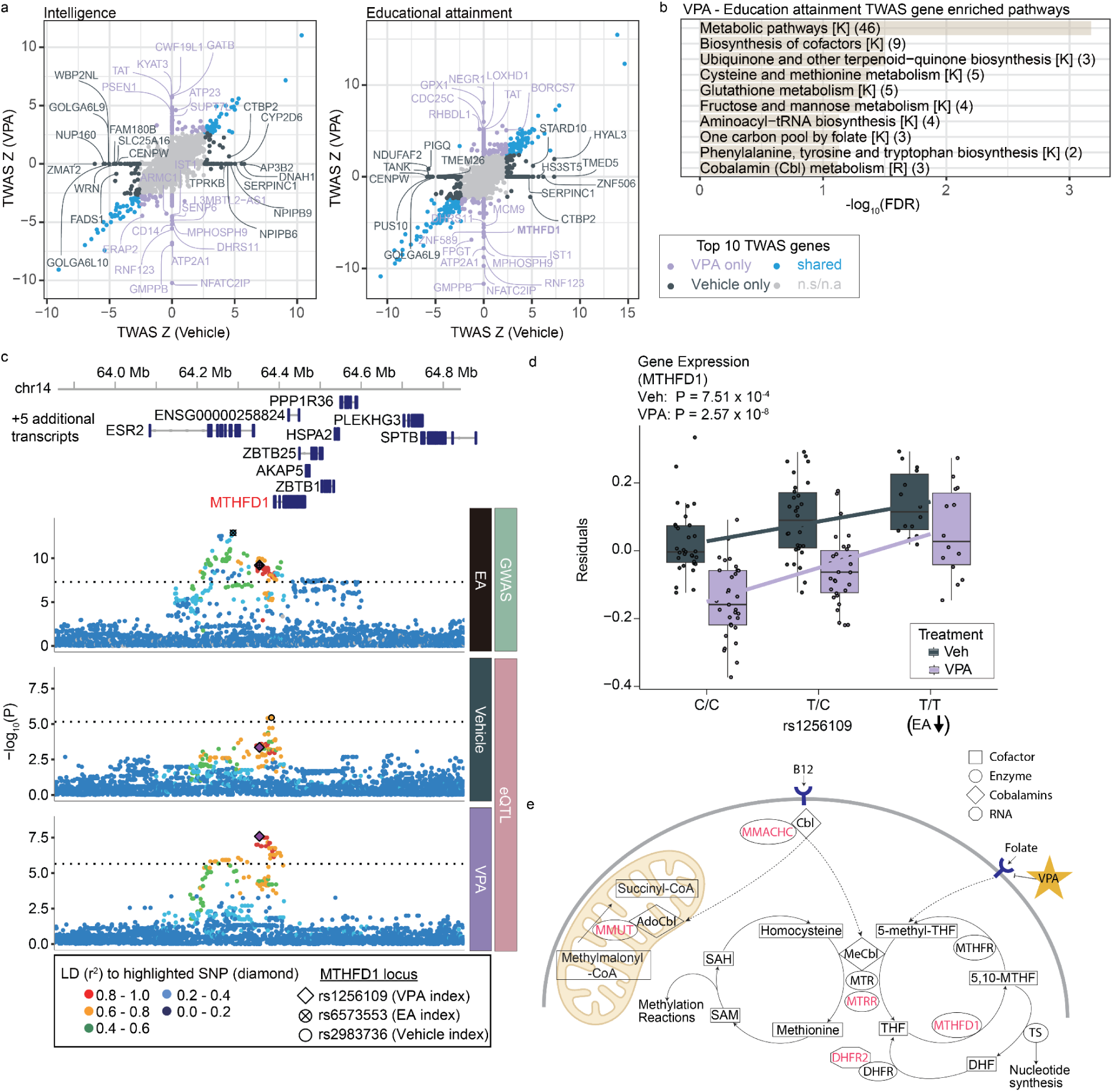
Drug-dependent TWAS identifies folate metabolism genes associated with cognitive ability. (a) Drug-dependent TWAS used to predict genes associated with intelligence^85^ (left) and educational attainment^79^ (right) during VPA exposure. Each dot indicates the TWAS gene Z score during VPA (y-axis) or vehicle treatment(x-axis). Genes without significant heritability were set to TWAS Z-scores of zero. (b) Pathways significantly enriched by genes associated with educational attainment after exposure to VPA. Numbers in parentheses indicate the intersection size, i.e., the number of DEGs overlapping with each pathway. K = KEGG, R = Reactome. (c) Regional association plot showing the co-localization of educational attainment GWAS (top panel) with a *MTHFD1*-modulating VPA eQTL (rs1256109). From top to bottom, the panels display genomic coordinates and gene models, *P* values for educational attainment GWAS, and P values for QTLs found in this study. (d) Allelic effects of rs1256109 on gene expression of *MTHFD1*. Dots indicate individual donors. The lower and upper hinges in box plots correspond to the first and third quartiles, and center lines correspond to the median of the residuals. (e) Abbreviated folate metabolism diagram, where VPA-dependent genes significantly associated with educational attainment are highlighted in red^87,88^. Star indicates a previously described site of VPA perturbation^11,12^.

Given the strong partitioned heritability enrichments and the greater number of QTLs detected with VPA, we focused on this exposure. We integrated QTLs with GWAS for educational attainment and cognitive ability, motivated by their large sample sizes and VPA’s connection to intellectual disability supported by our partitioned heritability findings (Fig. 1e). We detected hundreds of genes associated with differences in educational attainment using the TWAS approach, with many of the genes only detected using eQTL models trained in the VPA exposure (421 in vehicle, 384 in VPA, of which 191 genes are identified in both conditions) (Fig. 6a). We then sought replication for these findings using GWAS for intelligence. VPA TWAS Z scores were highly correlated (r = 0.62) and over 50% of significant VPA TWAS genes found in intelligence GWAS were also found in educational attainment (Supplementary Fig. 9c), which is expected given the high genetic correlation between both traits^79,85^ (*r_g_* = 0.73).

Next, we performed pathway enrichment analysis to functionally interpret the TWAS findings using the significant genes identified during treatment (Fig. 6b, Supplementary Fig. 9d, Supplementary Table 21). We found that genes associated with educational attainment during VPA exposure were significantly enriched for pathways related to folate and cobalamin (vitamin B12) metabolism (Fig. 6b). Among these genes (*DHFR2, MTFMT, MTHFD1, MMACHC, MTRR, MMUT*), a notable example is the *MTHFD1* gene. This key folate metabolism enzyme interconverts one-carbon derivatives of tetrahydrofolate and is significant only under VPA exposure. The VPA eQTL locus colocalized with a significant GWAS hit for educational attainment, where the rs1256109 variant C allele was associated with increased educational attainment and decreased *MTHFD1* expression (Fig. 6c-e).

Folate plays a crucial neurodevelopmental role in preventing neural tube defects which are exacerbated by exposure to VPA due to its interference with folate and related pathways^12^. Folate supplementation in mice rescues VPA induced teratogenicity^14^. Dietary folate supplementation in mothers taking antiepileptics, including VPA, is also associated with reduced autism diagnosis in the exposed children^30^. Finally, folic acid supplementation to VPA exposed organoids rescues some structural deficits^86^. These findings collectively demonstrate how genetic and drug-dependent regulatory analyses can identify genes and biological pathways associated with VPA-induced impacts on cognition, and suggest a treatment target that has already demonstrated efficacy in model systems and human populations.

## Discussion

In this study, we leveraged the inherent genetic variation in a population of genotyped, multi-ancestry human neural progenitor cells to analyze shifts in gene regulatory responses to clinically relevant exposures of VPA and Li. These drugs induce changes in chromatin accessibility and gene expression that lead to opposing effects on proliferation. Importantly, our work demonstrates that genetic variation alters the molecular response to drug exposures, serves as a foundation to better understand inter-individual variability in drug response, and suggests targetable treatment pathways to alleviate serious side effects. We noted a greater magnitude of these genetic effects following VPA exposure which may relate to lack of selective pressure on a synthetic molecule, in contrast to Li, which is a naturally occurring element where low-dose exposures are prevalent. Previous large scale pharmacogenomic studies in patient populations have recruited thousands of participants and yet identified only a few significant loci that are largely involved in pharmacokinetic effects, involved in drug metabolism, rather than pharmacodynamic effects, responses of brain cells to the drug^3^. Here, an alternative approach evaluated the impact of genetic variation on molecular response to drugs in a neural cell type using a “GxT in a dish” design that benefits from higher power through the use of controlled exposures and the study of molecular gene regulatory phenotypes. We were able to link the impact of these molecular impacts with clinical, behavioral, and brain structural outcomes using existing genome-wide association studies. Importantly, these analyses suggested that genetically mediated alterations in expression of folate metabolism enzymes during VPA exposure leads to inter-individual variability in educational attainment, a proxy for cognitive ability. Because VPA exposure leads to intellectual disability^4–6^, has known impacts on folate metabolism^12^, and folate supplementation alleviates the negative impacts of exposure in both mouse and human studies ^14,30^, our results demonstrate the utility of “GxT in a dish” approaches to both identify individuals who may be susceptible to the teratogenic impacts of VPA and provide further support to dietary folate supplementation during pregnancy when VPA is necessary.

Our work also supports a significant convergence of genetic and environmental factors contributing to neuropsychiatric disorder risk. We found both partitioned heritability enrichment of intelligence, educational attainment, and autism within VPA sensitive DARs as well as an enrichment of autism-associated genes within VPA DEGs. These results suggest that alterations in gene regulation occurring through either environmental exposures like VPA or through common or rare genetic variation can both result in increased risk for psychiatric disorders. We also found that drugs used to treat a disorder alter gene regulation within genes or regulatory elements that are associated with risk for the disorder. For instance, we detected a partitioned heritability enrichment of bipolar disorder genetic associations within Li DARs, and genes differentially expressed due to Li exposure were enriched in rare variants associated with bipolar disorder. Given that Li is a primary treatment for bipolar, these results suggest that this drug may be treating primary causal risk factors for the disorder.

A current question in the field of genetics of gene regulation is why eQTLs have different properties as compared to GWAS loci^70,71,89^. In contrast to GWAS loci, bulk eQTLs have been found to be enriched at promoter regions and affect genes less likely to be disease associated. Here, we observed that genes regulated by unique, context-dependent eQTLs are more distal to promoters and exhibit significantly lower LOEUF scores, indicating stronger negative selection. This suggests that context-dependent QTLs regulate genes that are functionally important and so under selective constraint, better matching disease GWAS loci and their role in disease risk, consistent with other emergent findings^69^. These context-dependent regulatory functions of variants would likely be missed under baseline conditions, unless very high sample sizes are acquired^90^.

This study highlights human neural progenitor cells as a valuable *in vitro* model for dissecting the intricate gene environment interactions that shape human brain development and influence complex neuropsychiatric traits. Our findings provide specific mechanistic insights into how clinically relevant environmental exposures converge with genetic predispositions. A limitation of the current study is the assessment of acute (48-hour) drug exposure effects. While informative, these findings may not fully capture the long-term or chronic effects of these drugs, which are often administered consistently over extended periods in clinical settings. Additionally, we studied the impacts of these drugs only in neural progenitor cells, which are relevant given that prenatal exposures to VPA lead to teratogenicity and Li impacts the proliferation of adult hippocampal progenitor cells^5,21^. Future research can build upon this foundation by exploring larger populations of primary hNPCs or iPSCs including differentiations to neuronal cell types, evaluating treatments for different durations, and assessing responses to a broader array of environmental insults or other pharmacological agents. Leveraging single cell multi-omics approaches will further refine our understanding of cell type specific responses in heterogeneous cultures like organoids. Ultimately, through consideration of exposure conditions, this work demonstrates a higher powered path to characterize gene x environment and gene x treatment interactions that are difficult to measure in human populations, potentially allowing the prioritization of individuals who will most benefit from treatment or use of complementary drugs to alleviate negative outcomes.

## Materials and methods

### Human neural progenitor cell (hNPC) culture

#### Ethics Statement for human tissue-derived cell lines

Human fetal brain tissue was obtained from voluntarily terminated pregnancies with the mother’s informed consent for the use of the tissue in research without compensation. All tissue was acquired and processed following the University of California, Los Angeles Gene and Cell Therapy Core’s institutional review board regulations.

#### Generation of hNPC lines

Fetal brain tissue with flat, sheet-like structure typical to the dorsal telencephalon was collected between 14 and 21 gestation weeks from donors presumed to be neurotypical^25,27,28^. Tissue was dissociated into single cells, cultured as neurospheres, and then transferred to 2-dimensional adherent cell culture on plates coated with Poly-L-Ornithine (Sigma P3655) and Fibronectin (Sigma F1141). Cells were maintained as proliferative neural progenitors^28^. NPC lines were cryopreserved and transferred to the University of North Carolina at Chapel Hill following expansion of 2-3 passages.

#### hNPC cell culture

Cryopreserved hNPC lines were thawed in batches of eight cell-lines per week. We pseudo-randomized each batch of cell-lines based on sex, tissue-donor gestation week, and passage number. We expanded cells for two weeks and then plated 400k hNPCs per well of a 6-well plate for each cell-line. We then exposed each cell-line to equal volumes of either 1mM Valproic acid sodium salt (P4543; Millipore Sigma) dissolved in water, 1.5mM LiCl (203637; Sigma-Aldrich) dissolved in water, or vehicle (hNPC media with PBS+0.1%BSA and DMSO). All exposures were prepared such that equal volumes were applied for each exposure condition, including a ‘balancer’ solution composed of culture media, PBS + 0.1% BSA, DMSO and water to standardize diluents across all exposures. Vehicle exposure conditions for each cell-line were identical to our previous study^29^. 48h after treatment, we lifted hNPCs (Accutase Thermo Fisher Scientific A1110501) for preparation of ATACseq and RNAseq libraries. Among these samples, we cultured 2-6 replicates for each of 6 randomly selected donors in distinct experimental weeks to evaluate technical variation throughout the experiment; one technical replicate from each donor–condition pair was selected for downstream analysis. When possible, we utilized media, additives, and treatments from the same manufacturer lots for the entire experiment to minimize potential batch effects. JMV conducted all hNPC cultures and treatments to reduce handling variance. Investigators were unblinded to hNPC line identity during cell culture or library preparation. No mycoplasma contamination was detected throughout the experiment (ATCC 30-1012K). hNPC media composition: Neurobasal A (Life Technologies 10888-022), 100 μg ml−1 primocin (Invivogen ant-pm-2), 10% BIT 9500 (Stemcell Technologies 09500), 1% glutamax (100x; Life Technologies 35050061), 1 μg ml^−1^ heparin (Sigma-Aldrich H3393-10KU), 20 ng ml^−1^ EGF/FGF (Life Technologies PHG0313/PHG0023), 2 ng ml^−1^ LIF (Life Technologies PHC9481) and 10 ng ml^−1^ PDGF (Life Technologies PHG1034).

### ATAC-seq library preparation

We prepared ATAC-seq libraries according to the “Omni-ATAC” protocol, using 50k hNPC nuclei following the vehicle, VPA, or LiCl exposure described above^91^. TDE1 enzyme (Illumina 20034198) tagmented these samples in a buffer of 20mM Tris-HCl pH 7.6, 10mM MgCl_2_, and 20% dimethyl formamide. Libraries were PCR amplified (NEB M0544S) for 5 cycles, and then further PCR amplified with additional cycles in a sample-specific manner determined by a qPCR side reaction to avoid overamplification. Across all samples, we used an average of 2 additional PCR cycles. During amplification, each sample was barcoded using Nextera-compatible unique dual-indexing primers (Illumina 20027213), and purified to remove fragments over 1000bp and primer dimers (Roche 07983298001). ATAC-seq library quality was verified in a subset of samples by detection of expected nucleosomal banding patterns on the Agilent 4150 TapeStation System (Agilent 5067-5584). We then pooled ∼70 libraries with unique barcodes for multiplexed sequencing. BDL generated all ATAC-seq libraries to minimize handling variation.

### RNA-seq library preparation

Total RNA-seq libraries were generated from the remaining hNPCs following the ATAC-seq library preparation described above, resulting in a mean of 1.1M hNPCs per sample. We isolated RNA for these preps using TriZol (Thermo 15-596-026) followed by RNA purification using RNeasy Mini Kits, and on-column DNAse digestion (Qiagen 74106, 79256). One individual, JTM, carried out RNA isolation to minimize handling variation. Quant-iT RNA assay kits (Thermo Q33140) quantified RNA isolates, which were then evaluated for quality via RNA integrity number (RIN). Average RIN across all samples was 9.86 (SD = 0.39). Finally, we depleted samples of ribosomal RNA (KAPA KR0934), and then prepared libraries with the stranded total RNA kit (KAPA KR0934).

### ATAC-seq and RNA-seq library sequencing

All libraries were sequenced at the New York Genome Center on the Illumina Novaseq platform. ATAC-seq libraries were sequenced to an average read depth of 49M paired-end (2x100) reads (SD = 13.6M). Total RNA-seq libraries were sequenced to an average read depth of 55M paired-end (2x100) reads (SD = 13M).

### Genotyping and imputation

Variants were directly genotyped with Illumina’s HumanOmni2.5 platform, using genomic DNA isolated from hNPCs with DNeasy Blood and Tissue Kit (QIAGEN 69504). Further variants were imputed on the University of Michigan’s imputation server using the TOPMed freeze 5 reference panel^92^ with minimac4 software^93^. Plink v1.981 software provided quality control, pre-processing, and filtering of SNPs based on Hardy-Weinberg equilibrium, minor allele frequency, individual missing genotype rates, and variant missing genotype rate (plink --hwe 1e-6 --maf 0.01 --mind 0.1 --geno 0.05).

### ATAC-seq data preprocessing

Quality control for all ATAC-seq libraries was implemented with FastQC (v0.11.9)^94^ and MultiQC (v1.7)^95^ software, before and after adapter trimming with BBMap (v38.98)^96^. The Burrows-Wheeler Alignment tool (BWA-MEM v0.7.17)^97^ aligned sequencing reads to the hg38 reference genome. To reduce mapping bias where reads overlapped bi-allelic SNPs, we remapped and removed duplicate reads with WASP (v0.3.4)^98^. Next, we removed unmapped or mitochondrial reads with Samtools (v1.16)^99^, and removed reads mapped to ENCODE-defined blacklist regions^100^ with bedtools (v2.3)^101^. Picard (v2.21.7, https://broadinstitute.github.io/picard/) and ataqv (v1.0.0)^102^ provided read metrics throughout preprocessing. We omitted samples with evidence of cross-contamination as evaluated by verifyBAMID (v1.1.3)^103^ when FREEMIX or CHIPMIX scores exceeded 0.02), with transcriptional start site enrichment below 5, and with anomalous short read/mononucleosomal read ratios (S/M < 1 or S/M > 7). Genotype data and BAM files from the same cell lines were used to identify sample swaps, also using verifyBAMID. High confidence swaps were adjusted to correct donor assignment (Original Identity-by-Descent (IBD) < .001 and corrected IBD > .98). Sex was not the primary driver of sample variation in the first two principal components (PCs) from our ATAC-seq data’s principal component analysis (PCA) (Supplementary Fig. 1). Since sex and Donor ID were correlated with other PCs, we included these variables in our quantitative trait loci (QTL) models and used residualized data after regressing out the first 10 PCs to control for their effects. Technical reproducibility of the ATAC-seq dataset is supported by the finding that pairwise correlations are significantly higher among technical replicates from the same hNPC donor and condition (“within donor”) than they are across distinct donors. A Welch’s two-sample t-test was performed for this evaluation (Supplementary Fig. 1).

### Chromatin accessibility peak calling

Chromatin accessibility peaks were called using CSAW’s (v1.28)^104^ *windowCounts* function using the following parameters: ext = mean length of ATAC-seq fragments, filter = 5 * sample number, spacing = 10, max.frag = 1500, pe=’both’, minq=20. After peak calling, windows within 100bp were merged via *mergeWindows*, and peak counts were normalized with conditional quantile normalization while accounting for GC-content^105^.

### RNA-seq data preprocessing

Like the ATAC-seq data, RNA-seq libraries were assessed for quality using FastQC and MultiQC. Next, we trimmed sequencing adapters and mapped reads to Ensembl v104 genes from the hg38 reference genome with STAR (v2.7.7a)^106^. Samples were omitted by the following criteria: (1) RNA integrity (RIN) scores below 7, (2) unique mapping rate below 80%, (3) mismatched read rate greater than 50%, (4) multi-mapping rates greater than 8%, (5) duplicated read rates greater than 30%, or (6) verifyBAMID FREEMIX or CHIPMIX scores greater than 0.02. We collapsed transcripts into a single gene model with collapse_annotation.py (https://github.com/broadinstitute/gtex-pipeline/tree/master/gene_model), and summarized gene-level reads with featureCounts from Rsubread (v2.8.2)^107^. Principal component analysis of RNA-seq samples showed that the first two principal components separated samples primarily based on their exposure condition, rather than by sex, RIN, or other biological and technical variables (Supplementary Fig. 1). However, since sex and donor ID were correlated with other PCs, they were included as variables in the subsequent quantitative trait locus (QTL) models. To account for this, the data used for these models was residualized after regressing out the first 10 PCs. Technical reproducibility of the RNA-seq dataset is supported by the finding that pairwise correlations are significantly higher among technical replicates from the same hNPC donor and condition (“within donor”) than they are across distinct donors. A Welch’s two-sample t-test was performed for this evaluation (Supplementary Fig. 1).

### Differential chromatin accessibility and gene expression analyses

DESeq2 (v1.44.0) assessed differential chromatin accessibility and gene expression as paired tests within hNPC donor after selecting pairs of vehicle and VPA- or LiCl-treated samples, prioritizing pairs cultured in the same plate that passed QC filtering. For differential chromatin accessibility, we used the following model:

Peak Accessibility ∼ Donor ID + treatment_condition.

For differential gene expression, we used the following model:

RNAseq Count ∼ RIN + Donor ID + treatment_condition.

In both cases, Donor ID and treatment condition were coded as factor variables. After estimating dispersion via shrunken log_2_ fold-change, we applied an FDR-adjusted *P* threshold of 0.1 to call DARs and DEGs. VPA and Li DARs were tested for significant bias in opened versus closed effect direction using “binom.test()” implemented in R.

### Chromatin feature enrichment analysis

All unique caPeaks across each drug condition were subset into DAR and non-DAR sets and overlapped with the 15 core chromHMM^40^ states defined in fetal brain male (E081). We assessed the significance of overlaps between our data and these features using a binomial test, checking if the number of annotated regions within the caPeak/eGene portion of the genome was statistically more frequent than random chance. Neighborhood enrichment scores were calculated based on Epigenomics Roadmap methodologies. To compare enrichment between two annotations, we used Fisher’s exact test. For multiple testing correction, both enrichment and Fisher’s test significance values were adjusted using the Benjamini-Hochberg procedure (FDR-adjusted *P* < 0.1).

### hNPC proliferation assays

We measured hNPC proliferation using Click-iT EdU-incorporation assays (Thermo Fisher Scientific C10337). Following two weeks of expansion as described above, we seeded 12.5k hNPCs per well of a 96-well plate including 4 technical replicates for each of the VPA and LiCl exposures, and 8 technical replicates for vehicle exposure, shuffling replicates and treatments across the plate to avoid potential plate-position effects. We exposed these hNPCs to the VPA, LiCl, or vehicle treatment for 48h. During the last 2h of this incubation, hNPCs were pulsed with 10μM EdU dissolved in media. Cells were then harvested and fixed with 4% paraformaldehyde in PBS for 10 minutes. Next, we followed manufacturer’s protocols to label total DNA content with FxCycle Far Red dye (Thermo Fisher Scientific F10347) and detect EdU-incorporation. Total DNA content and EdU signals were captured on the Attune NxT 96-well Flow Cytometer (Thermo Fisher Scientific), or via high content imaging on a Nikon Eclipse Ti2 microscope. FlowJo software (v10.7.1) filtered flow cytometry data for singlet signals, and we designated G0/G1, S, or G2/M cell-cycle populations using automated gating with FlowDensity software implemented in R^24,108^.

### Transcription factor binding site motif enrichment analysis

Using 841 predicted TFBS motifs from the JASPAR 2022 core vertebrate database ^43^, we assessed differential TFBS motif enrichment within VPA and LiCl DARs (Supplementary Table 4). This analysis focused on conserved regions greater than 20bp with 100-way phastCons scores greater than 0.4 ^109^. Using the top 2,000 DARs either opened or closed by treatment (|log_2_FC|, FDR-adjusted *P* < 0.1), we performed logistic regression using the following model:

glm(TFBS ∼ peaktype + peakwidth + percent-conservedBP, family = ‘binomial’)

where “TFBS” is a binary outcome reflecting whether each chromatin accessibility peak overlaps with a given TFBS motif, “peaktype” is a binary indicator of whether a peak is an open or closed DAR for each treatment condition, “peakwidth” encodes the size of the peak’s genomic footprint in base-pairs, and “percent-conservedBP” encodes the percentage of conserved base-pairs within a peak (conservedBP / peakwidth). This approach identifies TFBS motifs observed more often in opened versus closed DARs while accounting for peak size and conserved sequences. Estimated effects of “peaktype” on TFBS motif presence with FDR-adjusted *P* < 0.1 were considered statistically significant.

### Partitioned Heritability

We used stratified linkage disequilibrium score regression to evaluate the enrichment of neuropsychiatric, cognitive, and structural brain-trait heritability within DARs^51,110^. Publicly available summary statistics from various brain-trait GWAS (Supplementary Table 16) were downloaded and filtered for HapMap3 SNPS using LDSC’s munge_sumstats.py script. We selected European population summary statistics when associations for multiple ancestries were available to better match European population LD-scores from the 1000 Genomes Project (phase3), which were pre-computed and downloaded from https://data.broadinstitute.org/alkesgroup/LDSCORE/eur_w_ld_chr.tar.bz2 ^111^. Heritability enrichments for LiCl/VPA DARs (those with FDR-adjusted *P* < 0.1) were then calculated by including non-DARs using the baseline model (v1.2). One-tailed heritability enrichment *P*-values calculated from each DAR annotation’s Z coefficient were then FDR-adjusted separately for LiCl and VPA DARs, setting the significance thresholded at FDR-adjusted *P* < 0.1.

### Gene Ontology/ Pathway Enrichment

The top 1000 up-regulated or down-regulated LiCl or VPA DEGs ranked by FDR-adjusted *P* passing a significance threshold of *P* < 0.1 were used as input for gene ontology enrichment. These DEGs were evaluated against a background composed of all other genes evaluated in DEG analysis after filtering for protein-coding genes located outside the MHC region (chr6:28,510,120-33,480,577). Enrichment tests for Gene Ontology terms from the KEGG and REACTOME pathways^112,113^ were run using g:profiler (v0.2.3)^114^, with significant enrichment determined at a threshold of FDR-adjusted *P* < 0.1. For gene ontology enrichment tests of genes encoding transcription factors with TFBS motifs found to enriched within open or closed DARs (Supplementary Table 5), we used a background of all protein-coding TF genes from the JASPAR 2022 core vertebrate database^43^.

### Gene Set Enrichment Tests

Fisher exact tests implemented in R evaluated overlap between upregulated or downregulated DEGs and gene sets from either VPA/LiCl response studies in similar cell types^35,60,61^ or neuropsychiatric risk genes implicated by exome sequencing studies^62,115^. *P*-values from these tests were FDR-adjusted for multiple comparisons separately for VPA and LiCl DEGs. Positive odds ratios denote a higher degree of overlap between DEGs and the gene sets from other studies than expected.

### Chromatin accessibility quantitative trait loci (caQTL) mapping

We performed caQTL analyses for each condition (vehicle, VPA, and LiCl) using the RASQUAL method^116^. This joint model accounts for both genetic differences between individuals and allele-specific effects within them. We incorporated CQN normalization factors as sample-specific offsets and included 10 count-based principal components (PCs) and 10 multi-dimensional scaling (MDS) genotype components as covariates to control for technical variations and population structure, respectively. Variants were tested by the following criteria: (1) located in an accessible region or within ±25 kb (2) having a MAF ≥ 1% (3) Hardy-Weinberg equilibrium <0.000001 (4) imputation quality ≥ 0.3; and (5) at least two individuals with either a minor homozygous or heterozygous allele. To account for multiple comparisons, we first employed eigenMT to estimate the number of independent statistical tests conducted in the cis region of each feature^117^. We then applied Bonferroni correction to acquire adjusted *P-*values. RASQUAL was also run with the ’--random-permutation’ setting to generate empirical null *P* values by permuting sample labels. We then put these permuted *P*-values through the same eigenMT multiple testing correction process previously described. Observed adjusted *P* values were then compared to the empirical null distribution to identify QTLs with FDR adjusted *P* < 0.1 Due to RASQUAL’s limitations, we were unable to perform true conditional analysis. Instead, to identify distinct signals, we iteratively used an LD threshold of r^2^ < 0.2 until no further significant caQTLs were found.

For response caQTL interaction analyses between vehicle and each treatment (VPA or LiCl), we used the R package lme4 to evaluate the following linear mixed model for all index caQTL with main effects (n_caSNP_ = 32,343 (VPA–vehicle pairs), 34,024 (LiCl–vehicle pairs):

𝑎𝑑𝑗𝐶𝑜𝑢𝑛𝑡𝑠 = 𝑆𝑁𝑃 + 𝑐𝑜𝑛𝑑𝑖𝑡𝑖𝑜𝑛 + 𝑐𝑜𝑛𝑑𝑖𝑡𝑖𝑜𝑛: 𝑆𝑁𝑃 + 𝑐𝑜𝑣𝑎𝑟𝑖𝑎𝑡𝑒𝑠 + (1|𝐷𝑜𝑛𝑜𝑟)

The “condition” variable was set to 0 for vehicle and 1 for the drug condition and the first 10 count-based PCs served as covariates. Significance was then determined by comparing the model’s fit with and without the “condition:SNP” interaction term. Multiple testing correction was then performed using the Benjamini–Hochberg procedure (FDR-adjusted *P* < 0.1).

### Gene expression quantitative trait loci (eQTL) mapping

Linear mixed models estimated genetic effects on gene expression were estimated within each of the vehicle, LiCl, VPA conditions using limix_qtl^118,119^. We paired protein-coding or lncRNA genes with 10 or more normalized counts in 10% or more samples, with SNPs located inside or within 1Mb of gene bodies with MAF ≥ 1%, Hardy–Weinberg equilibrium < 0.000001, imputation quality ≥ 0.3, and where at least two samples possess the minor allele. These gene-SNP pairs were evaluated using the model: 𝑎𝑑𝑗𝐸 = 𝑆𝑁𝑃 + ε.

In this model, 𝑎𝑑𝑗𝐸 represents VST-normalized gene read count residuals after correction with 10 principal components of gene expression variance, SNP represents the 0/1/2 genotype of the tested allele, and ε represents an error term with cov(*ɛ*) = (σ^2^_u_*u_K_*+ σ^2^_e_*I*), where *u_K_* is a kinship matrix generated by GENESIS v2.14.1^120^ using 10 PCs of genotypic variance, σ^2^_u_ is variance from genetic relatedness, and σ^2^_e_ is the variance due to random noise. We corrected eQTL *P*-values for multiple testing at the gene level using 1000 permutation tests in limix_qtl, followed by Benjamini-Hochberg-FDR at the global level. SNP-Gene pairs reaching an FDR-adjusted *P* less than 0.1 were considered as statistically significant eQTLs (eSNP-eGene pairs).

We identified response eQTLs within significant index eQTLs with main effects through condition by genotype interaction analysis in limix_qtl for either vehicle and LiCl samples (2,716 pairs), or vehicle and VPA (2,750 pairs) using the following model:

*adjE = SNP + condition + condition:SNP + ε ,*

where *SNP* and *ε* were coded as described above, and *condition* was coded as 0 for vehicle or 1 for either LiCl or VPA treatment. Significant SNP:condition interaction effects were defined as those with FDR-adjusted *P* were less than 0.1.

### Evaluating specificity of eSNPs/caSNP condition

To assess specificity of eSNP/caSNP discovery, we compared across each of the exposure conditions (vehicle, VPA, LiCl) as well as WNT stimulated conditions (CHIR and WNT3A) from our previous study^29^. The analysis used eSNP/caSNPs identified in the Li/VPA conditions as the reference pool. This reference pool of SNPs was then compared against each other and against the SNPs identified in the WNT-stimulated conditions. All condition pairs were tested for overlapping loci, defined as lead ca/eQTLs in LD r^2^ > 0.8. An overlapping eSNP required the same associated gene, while an overlapping caSNP required overlapping peak calls. Those results were then binned into three discovery categories: (1) Unique, discovered in a single condition; (2) Multiple, discovered in 2-4 conditions; and (3) Shared, common across all conditions. We then investigated differences across discovery categories for eGenes in two ways: (1) determining the absolute distance from the TSS for each gene and (2) comparing intolerance to loss-of-function variation using pLOEUF (predicted loss-of-function observed/expected upper bound fraction) scores from gnomAD (v2.1.1). Significance between each discovery category was then determined by Wilcoxon rank sum test for both distance from TSS and pLOEUF score comparisons.

### Identifying eSNPs/caSNPs found in GWAS

We conducted a three-step approach to assess co-localization between our identified ca/e-QTL variants and brain-related GWAS signals. First, we extracted all GWAS lead SNP-ca/eQTL pairs (*P* < 5 × 10^−8^) within 1 Mb of each other. Next, we selected GWAS-QTL pairs where index SNPs were in strong LD (*r^2^* > 0.6) in either the 1000 Genomes European population or our study’s samples. To capture potential undefined secondary GWAS signals, we also included pairs where a non-index, genome-wide significant GWAS SNP (*P* < 5 ×10^−8^) was in LD with a ca/eQTL index SNP. Finally, we then tested for co-localization using eCAVIAR, calculating the SNP-level co-localization posterior probability (CLPP) for each SNP that satisfied the conditions of the preceding two steps. CLPP was estimated for SNPs with *P*<0.05 in both GWAS and QTL data. Sites were considered co-localized if any locus had a CLPP greater than 1% and an *r^2^* between a causal SNP and at least one genome-wide SNP in the region greater than 0.8. Understanding that false positives for QTL condition specificity could arise due to insufficient power from limited sample size, we performed a repeated eCAVIAR analysis. This was done to retain potentially shared eGene/caPeak–GWAS pairs that, while not meeting the FDR-adjusted *P* < 0.1, did pass the raw significance threshold (*P* < 5 x 10^−6^) for QTL mapping under a specific condition and demonstrated shared caPeak/eGene–GWAS pairs under a different condition. Ssimp software (https://github.com/zkutalik/ssimp_software/tree/master/)^121,122^ was used for imputation of summary statistics for rs57533386 from EA2022 GWAS summary statistics^79^ and the 1KG reference panel.

### TWAS

Transcriptome-wide association on autosomal chromosomes was conducted using FUSION software (http://gusevlab.org/projects/fusion/)^81^. We used the same gene expression matrices as in eQTL mapping, consisting of residualized VST normalized expression values. SNPs within 1 Mb of the gene body were considered, but restricted to those overlapping with the European (EUR) reference panel to construct the LD reference panel. In our previous study where we analyzed nearly identical donor samples, we observed large overlaps in TWAS results between the EUR and the study-specific populations^27^. Thus, we only analyzed our own LD panel. After filtering for genes with for genes with heritability *P* < 0.01, we estimated gene expression weights in each condition using multiple models (BLUP, BSLMM, LSSO, top SNPs, and Elastic Net) implemented in FUSION. The predicted gene expression values with the highest R² were selected for TWAS and preprocessed GWAS summary statistics prepared for partitioned heritability analysis were utilized. Significant associations were defined as those with FDR-adjusted *P* < 0.1 within each GWAS-condition. Pathway enrichment analysis on TWAS genes was performed using g:profiler^114^ targeting the Reactome and KEGG databases^112,113^.

## Data Availability

All data and metadata generated for this study will be available via dbGaP; acccession number TBD. All code used in analyses and data including full summary statistics except that which is provided in supplementary tables will be available on bitbucket.

https://bitbucket.org/steinlabunc/clinical_rqtls/

## Acknowledgements

This study was supported by grants from the National Institute of Mental Health (NIMH) (R01MH118349, R01MH120125 and R01MH121433). J.M.V. and B.D.L. were supported, in part, by National Institutes of Health (NIH) T32 training grants (T32GM135123 and T32GM067553, respectively). We gratefully acknowledge technical support from UNC Research Core facilities, which are supported by University Cancer Research Fund Comprehensive Cancer Center Core Support Grant P30-CA016086. The UNC Flow Cytometry Core Facility is supported, in part, by North Carolina Biotech Center Institutional Support Grant 2017-IDG-1025 and NIH 1UM2AI30836-01. The funders had no role in study design, data collection and analysis, decision to publish or preparation of the manuscript. We thank A. Beltran, S. G. Molina and M. Calabrese for access to instruments. We would like to acknowledge the use of bioRender (www.biorender.com) templates to make the cartoons in Figures 1 and 2.

## Author contributions

J.M.V., B.D.L. and J.L.S. designed the study. J.M.V. maintained hNPC cultures and applied drugs. B.D.L. and J.M.W. performed proliferation assays. B.D.L. generated ATAC-seq libraries. J.T.M. isolated RNA samples. J.M.V. and B.D.L. processed and prepared ATAC-seq and RNA-seq data. M.I.L. advised on statistical methods. J.M.V., B.D.L., and N.M. performed quality control and analyses on sequencing data. J.M.V., N.M., B.D.L. and J.L.S. prepared the initial draft of the manuscript. All authors read and approved the final manuscript.

## Resource availability

### Lead contact

Correspondence and requests for materials should be addressed to Jason L. Stein.

### Materials availability

Data and metadata generated for this study will be available via dbGaP; acccession number TBD

### Data and code availability

All code used in analyses and data including full summary statistics except that which is provided in supplementary tables will be available at https://bitbucket.org/steinlabunc/clinical_rqtls/.

## Supplemental information

Supplemental Figs. S1 to S9

Tables S1 to S21

