## Supplementary Figures for "Assessing molecular gene by treatment interactions using a population of neural progenitors exposed to valproic acid and lithium"

### Extended Data - Li/VPA-QTL MS


Supplementary Figures

- Supplementary Figure 1 | Assessing Technical Reproducibility and Principal Component Analysis in Gene Expression and Chromatin Accessibility Studies.
- Supplementary Figure 2 | Pathway enrichment analysis results.
- Supplementary Figure 3 | Differential HDAC expressions between VPA and Vehicle
- Supplementary Figure 4 | Identification of putative gene regulatory elements through peak-gene correlation.
- Supplementary Fig. 5 | QTL effect sizes show strong correlation across conditions.
- Supplementary Fig. 6 | Responsive caQTL after VPA exposure colocalizes with an eQTL for STAT4.
- Supplementary Fig. 7 | The colocalization of eQTLs/caQTLs with brain-related GWAS phenotypes tested by eCAVIAR.
- Supplementary Fig. 8 | Colocalization of SUPT7L associated r-caQTL with average cortical thickness.
- Supplementary Figure 9 | TWAS implicates condition-dependent gene-trait associations.

Supplementary Tables

- Supplementary Table 1: DARs
- Supplementary Table 2: chromHMM Enrichment values
- Supplementary Table 3: chromHMM Fisher values
- Supplementary Table 4: TF enrichments
- Supplementary Table 5: Pathway enrichments
- Supplementary Table 6: LDSC
- Supplementary Table 7: DEGs
- Supplementary Table 8: Peak-gene correlation
- Supplementary Table 9: caQTLs
- Supplementary Table 10: eQTLs
- Supplementary Table 11: r-caQTLs
- Supplementary Table 12: r-eQTLs
- Supplementary Table 13: caQTL-eQTL overlaps
- Supplementary Table 14: r-QTL context discovered
- Supplementary Table 15: eQTL context discovered and pLOEUF score
- Supplementary Table 16: GWAS list
- Supplementary Table 17: caQTL GWAS overlaps
- Supplementary Table 18: eQTL GWAS overlaps
- Supplementary Table 19: caQTL eCAVIAR colocalizations
- Supplementary Table 20: eQTL eCAVIAR colocalizations
- Supplementary Table 21: TWAS results

### Supplementary Figures

#### Supplementary Figure 1 | Assessing Technical Reproducibility and Principal Component Analysis in Gene Expression and Chromatin Accessibility Studies.

Technical reproducibility of measurements calculated by pairwise correlations of normalized sequencing counts. These correlations were derived from either open chromatin peaks (ATAC-seq, a) or gene expression levels (RNA-seq, b) across technical replicates. Robust reproducibility is indicated by higher correlations within a single donor's replicates compared to correlations across different donors. Violin plots illustrate the distribution of Pearson correlation coefficients, comparing "across donor" pairs (genotypically distinct individuals) with "within donor" pairs (technical replicates from the same donor, cultured at different times). The top and bottom horizontal lines mark the interquartile range, and the middle bar indicates the median. Fisher's Z transformation followed by a two-sided Welch two-sample t-test were used to calculate p-values, which report significant differences between the "across donor" and "within donor" correlations. The x-axis shows the total number of cell lines and pairwise correlations (n) for each experimental condition. Principal component analysis (PCA) was performed on ATAC-seq (c, d) and RNA-seq (e, f) data counts following variance stabilizing transformation (VST). Samples are annotated by experimental condition and biological sex. Correlation matrices of ATAC-seq (g) and RNA-seq (h) principal components (1-10) for each condition (VPA or Li) incorporating technical (RIN) and biological (donor and sex) variables. Linear regression was applied to remove the effects of principal components 1-10. The resulting residualized sequencing count data was subsequently utilized as input for quantitative trait loci (QTL) modeling, accounting for both measured and unmeasured confounding factors.

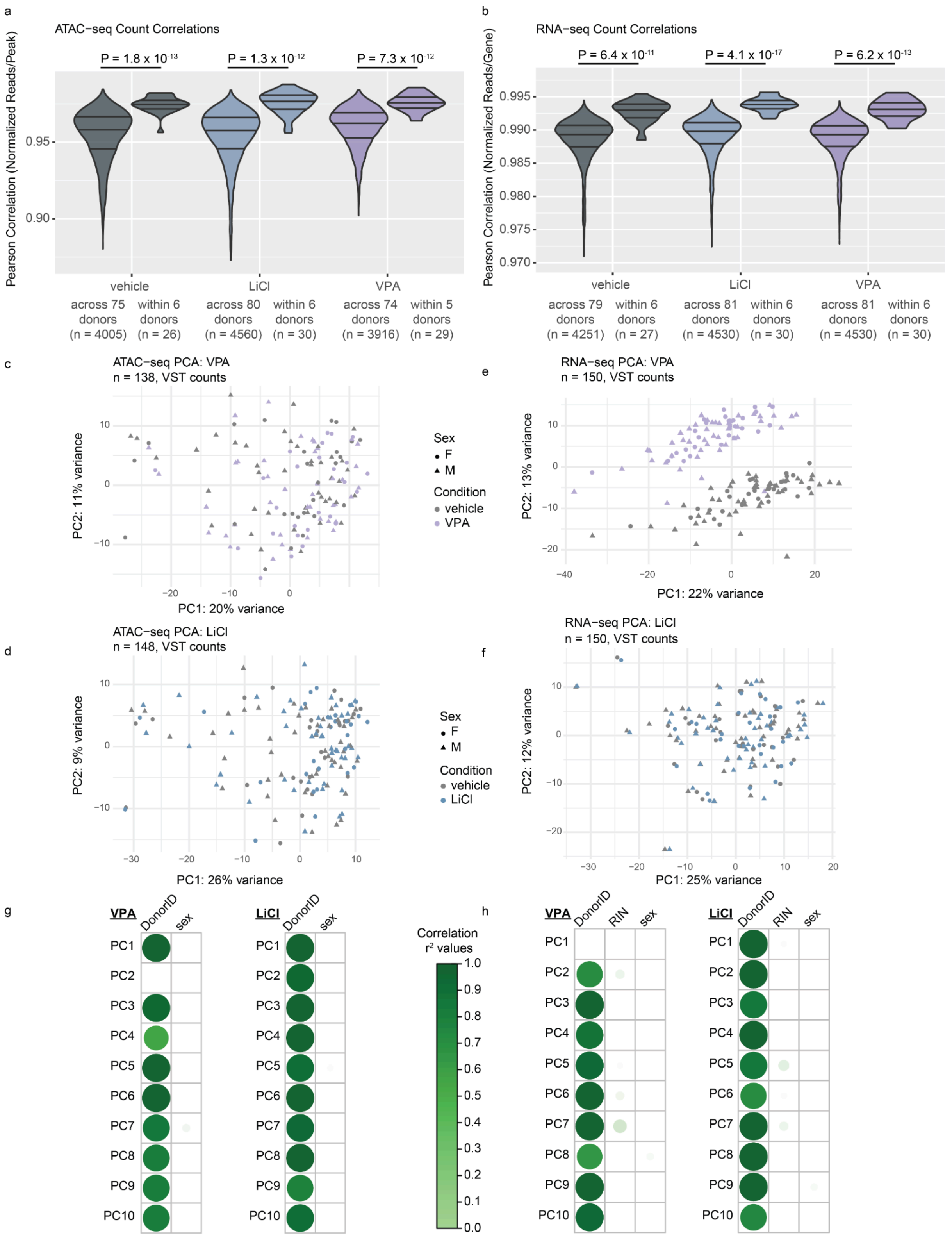

a
ATAC−seq Count Correlations
P = 1.8 x 10-13
P = 1.3 x 10-12
P = 7.3 x 10-12
Pearson Correlation (Normalized Reads/Peak)
vehicle
LiCl
VPA
across 75 donors (n = 4005)
within 6 donors (n = 26)
across 80 donors (n = 4560)
within 6 donors (n = 30)
across 74 donors (n = 3916)
within 5 donors (n = 29)
b
RNA−seq Count Correlations
P = 6.4 x 10-11
P = 4.1 x 10-17
P = 6.2 x 10-13
Pearson Correlation (Normalized Reads/Gene)
across 79 donors (n = 4251)
within 6 donors (n = 27)
across 81 donors (n = 4530)
within 6 donors (n = 30)
across 81 donors (n = 4530)
within 6 donors (n = 30)
c
ATAC−seq PCA: VPA
n = 138, VST counts
PC2: 11% variance
PC1: 20% variance
Sex
F
M
Condition
vehicle
VPA
e
RNA−seq PCA: VPA
n = 150, VST counts
PC2: 13% variance
PC1: 22% variance
d
ATAC−seq PCA: LiCl
n = 148, VST counts
PC2: 9% variance
PC1: 26% variance
Sex
F
M
Condition
vehicle
LiCl
f
RNA−seq PCA: LiCl
n = 150, VST counts
PC2: 12% variance
PC1: 25% variance
g
VPA
DonorID
sex
LiCl
DonorID
sex
Correlation r² values
h
VPA
DonorID
RIN
sex
LiCl
DonorID
RIN
sex

### Supplementary Figure 2 | Pathway enrichment analysis results.

Top 10 pathways or Gene Ontology (except for GO:CC) enriched in transcription factors found in 2,000 opened or closed peaks due to VPA (left) or Li (right) exposures. Red line indicates FDR = 0.1. The full list can be found in Supplementary Table 4.

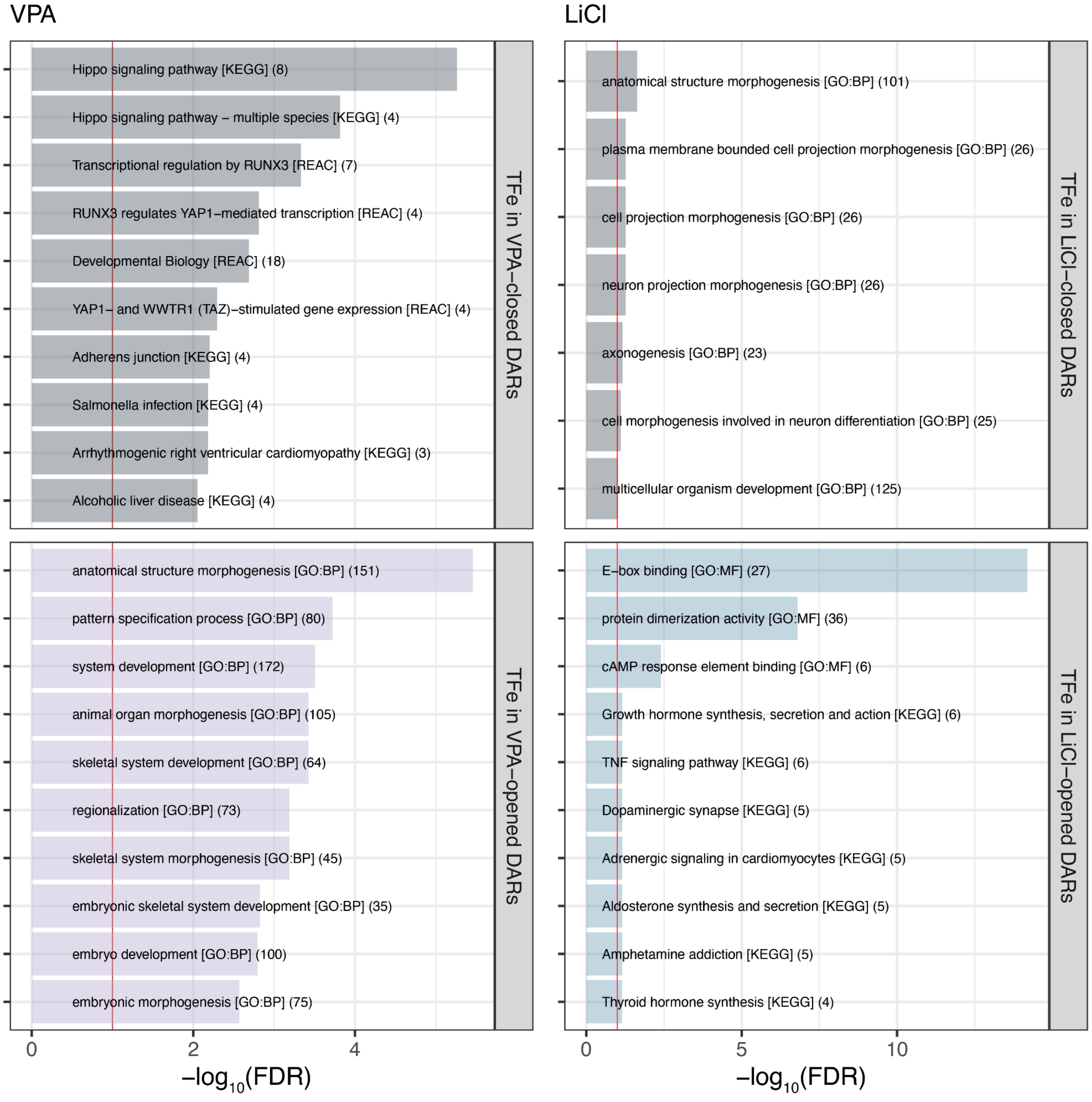

### Supplementary Figure 3 | Differential HDAC expressions between VPA and Vehicle

VPA directly binds to inhibit HDAC class I and IIa proteins. Here, we show that most HDACs, other than HDAC7 and 10, are upregulated following VPA exposure, perhaps as a compensatory mechanism following protein inhibition.

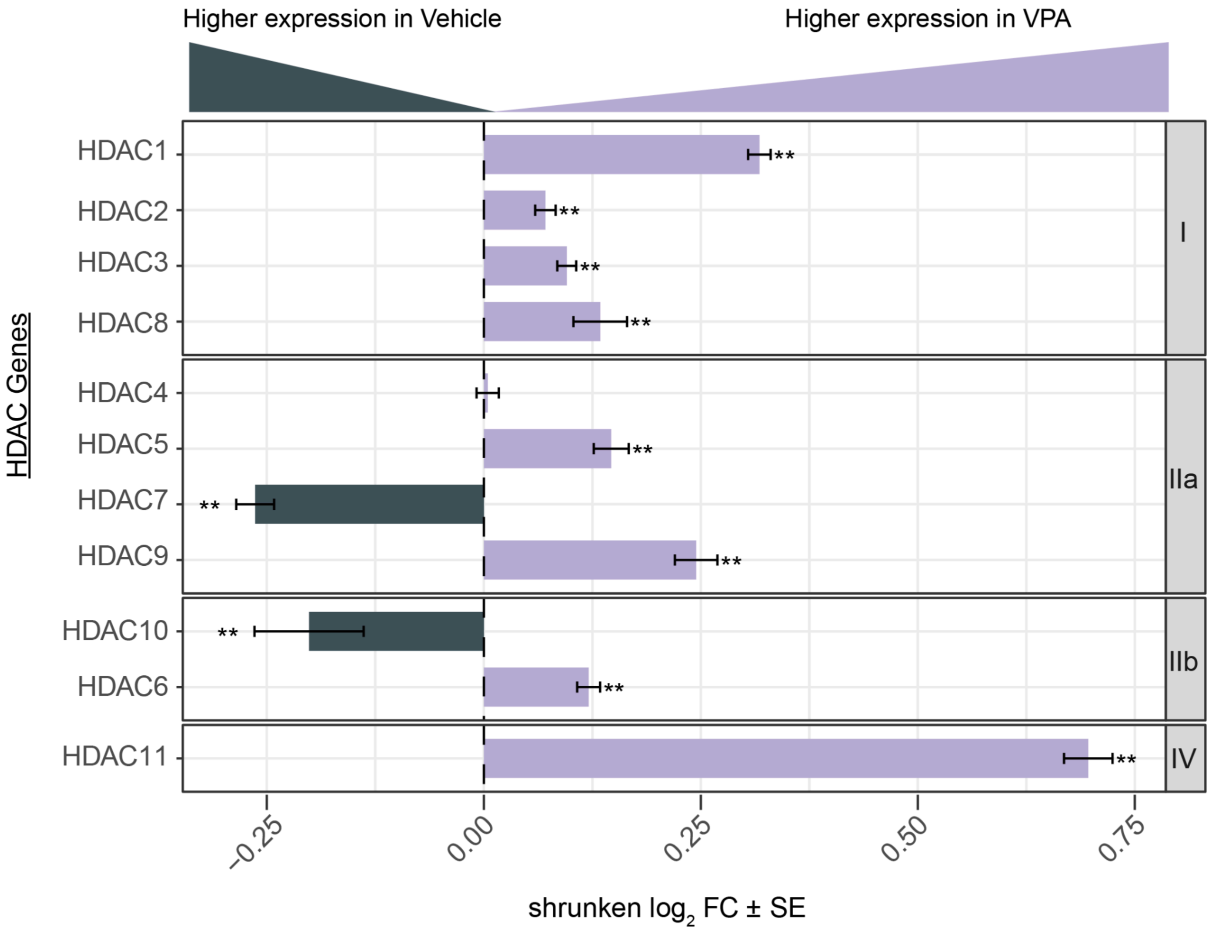

#### Supplementary Figure 4 | Identification of putative gene regulatory elements through peak-gene correlation.

Regulatory peaks putatively influencing gene expression were identified through correlation analysis of expression and chromatin accessibility. Panels (a-c) display the distribution of absolute peak distance to the transcription start site (TSS) for correlated genes under Vehicle, VPA, and Li, respectively. (d) quantifies significant correlated peak-gene pairs. A total of 12,516 peak-gene pairs were uniquely detected under treatment conditions, suggesting drug-dependent regulatory relationships. (e) Regulatory peaks correlated with *NRXN3* gene expression during Vehicle or Li exposure. (f) Pathway enrichment of genes with correlated peak pairs only under VPA (top) or Li (bottom), but not Vehicle.

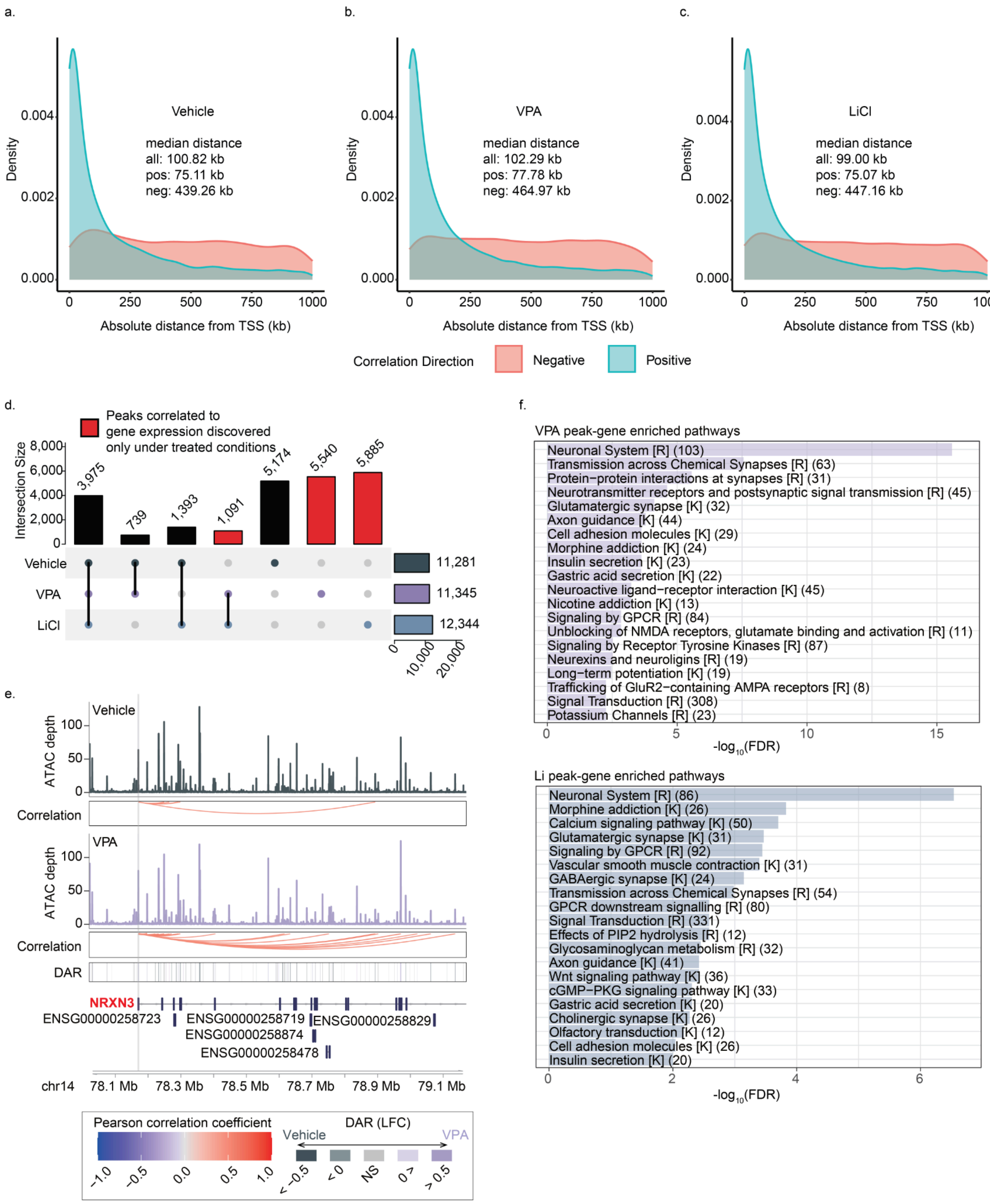
a.
Vehicle
median distance
all: 100.82 kb
pos: 75.11 kb
neg: 439.26 kb
Density
Absolute distance from TSS (kb)
b.
VPA
median distance
all: 102.29 kb
pos: 77.78 kb
neg: 464.97 kb
Density
Absolute distance from TSS (kb)
c.
LiCl
median distance
all: 99.00 kb
pos: 75.07 kb
neg: 447.16 kb
Density
Absolute distance from TSS (kb)
Correlation Direction
Negative
Positive
d.
Peaks correlated to gene expression discovered only under treated conditions
Intersection Size
3,975
739
1,393
1,091
5,174
5,540
5,885
Vehicle
VPA
LiCl
11,281
11,345
12,344
e.
ATAC depth
Vehicle
Correlation
VPA
Correlation
DAR
NRXN3
ENSG00000258723
ENSG00000258719
ENSG00000258829
ENSG00000258874
ENSG00000258478
chr14
78.1 Mb
78.3 Mb
78.5 Mb
78.7 Mb
78.9 Mb
79.1 Mb
Pearson correlation coefficient
DAR (LFC)
Vehicle
VPA
< −0.5
< 0
NS
0 >
> 0.5
f.
VPA peak-gene enriched pathways
Neuronal System [R] (103)
Transmission across Chemical Synapses [R] (63)
Protein−protein interactions at synapses [R] (31)
Neurotransmitter receptors and postsynaptic signal transmission [R] (45)
Glutamatergic synapse [K] (32)
Axon guidance [K] (44)
Cell adhesion molecules [K] (29)
Morphine addiction [K] (24)
Insulin secretion [K] (23)
Gastric acid secretion [K] (22)
Neuroactive ligand−receptor interaction [K] (45)
Nicotine addiction [K] (13)
Signaling by GPCR [R] (84)
Unblocking of NMDA receptors, glutamate binding and activation [R] (11)
Signaling by Receptor Tyrosine Kinases [R] (87)
Neurexins and neuroligins [R] (19)
Long−term potentiation [K] (19)
Trafficking of GluR2−containing AMPA receptors [R] (8)
Signal Transduction [R] (308)
Potassium Channels [R] (23)
-log10(FDR)
Li peak-gene enriched pathways
Neuronal System [R] (86)
Morphine addiction [K] (26)
Calcium signaling pathway [K] (50)
Glutamatergic synapse [K] (31)
Signaling by GPCR [R] (92)
Vascular smooth muscle contraction [K] (31)
GABAergic synapse [K] (24)
Transmission across Chemical Synapses [R] (54)
GPCR downstream signalling [R] (80)
Signal Transduction [R] (331)
Effects of PIP2 hydrolysis [R] (12)
Glycosaminoglycan metabolism [R] (32)
Axon guidance [K] (41)
Wnt signaling pathway [K] (36)
cGMP−PKG signaling pathway [K] (33)
Gastric acid secretion [K] (20)
Cholinergic synapse [K] (26)
Olfactory transduction [K] (12)
Cell adhesion molecules [K] (26)
Insulin secretion [K] (20)
-log10(FDR)

### Supplementary Fig. 5 | QTL effect sizes show strong correlation across conditions.

(a-d) Beta values of QTLs identified in this study. Comparison of effect sizes for significant index caQTLs (a, c) and eQTLs (b, d) between treated and vehicle conditions (VPA or Li in (a, b) and (c, d) respectively). Each dot represents one index SNP. Red dots show significant interaction effects (r-QTLs). Two-sided Pearson correlation (r) and P-values were estimated per condition pairing. The black regression line is indicated for all significant index QTLs, while the red regression line and its 95% confidence interval are shown for r-QTLs. A dotted line indicating y=x is also included for reference. r-QTLs exhibited a lower correlation compared to non-r-QTLs, as expected. Note, some SNPs lacked a beta value from the baselines model due to low expression and so are not plotted. $n_{VPA\text{-}caQTL}$ = 9,094, $n_{VPA\text{-}rcaQTL}$ = 779 , $n_{Li\text{-}caQTL}$ = 10,693, $n_{Li\text{-}rcaQTL}$ = 15 (a, b); $n_{VPA\text{-}eQTL}$ = 605, $n_{VPA\text{-}reQTL}$ = 147, $n_{Li\text{-}eQTL}$ = 659, $n_{Li\text{-}reQTL}$ = 15 (c, d).

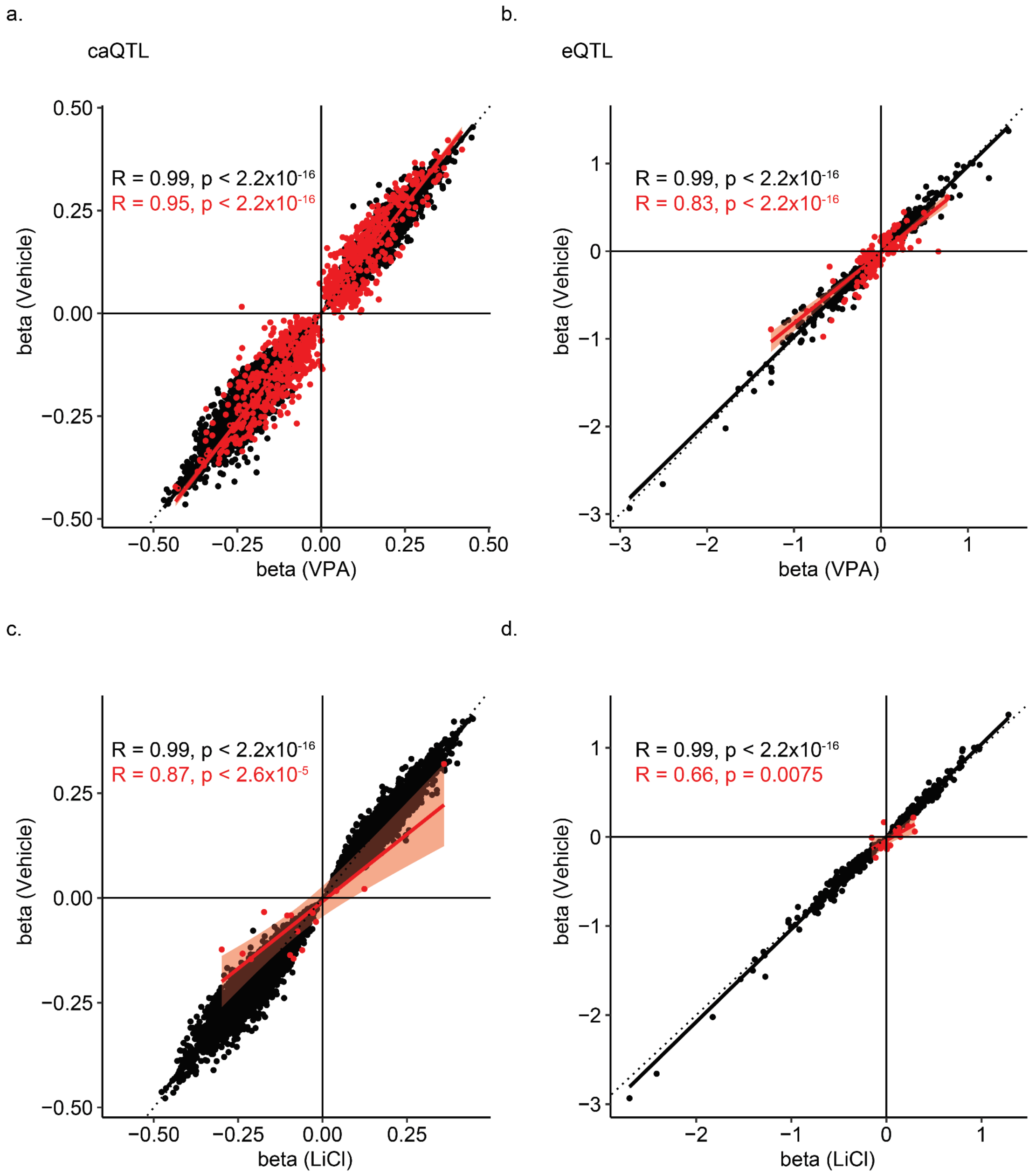

### Supplementary Fig. 6 | Responsive caQTL after VPA exposure colocalizes with an eQTL for *STAT4*.

(a) Boxplot showing an increase in chromatin accessibility associated with rs2356354-C. An interaction effect between vehicle and VPA conditions significantly mutes the effect. $n_{vehicle}$ = 76, $n_{VPA}$ = 75. (b) Boxplot showing increased *STAT4* gene expression by rs1454749-C. nvehicle = 76, nVPA = 78. P values for eQTLs were estimated by a linear mixed model using a two-sided test. caQTL P values were obtained using a likelihood-ratio test in RASQUAL (Methods). QTLs were adjusted for multiple testing correction by the Benjamini–Hochberg procedure. The lower and upper hinges in box plots correspond to the first and third quartiles, and center lines correspond to the median of the residuals.

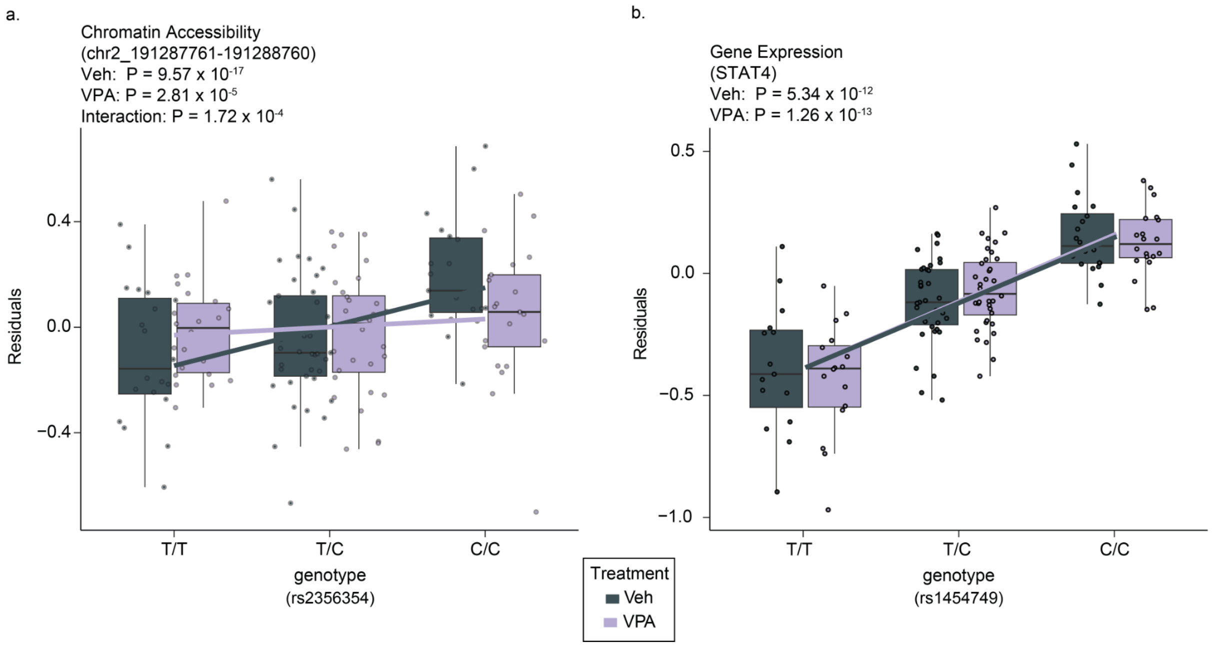

### Supplementary Fig. 7 | The colocalization of eQTLs/caQTLs with brain-related GWAS phenotypes tested by eCAVIAR.

(a) The count of caPeaks (left) and eGenes (right) that overlap brain-related GWAS loci, as identified by eCAVIAR shown for vehicle, VPA, Li, and drug-dependent caPeaks/eGenes. (b) The cumulative total of unique colocalized eGenes/caPeaks following eCAVIAR analysis. (c) Shared versus distinct GWAS-feature pair colocalizations across conditions. Stimulated conditions notably increase brain-trait associated genes by 134.8% and peaks by 49.7%.

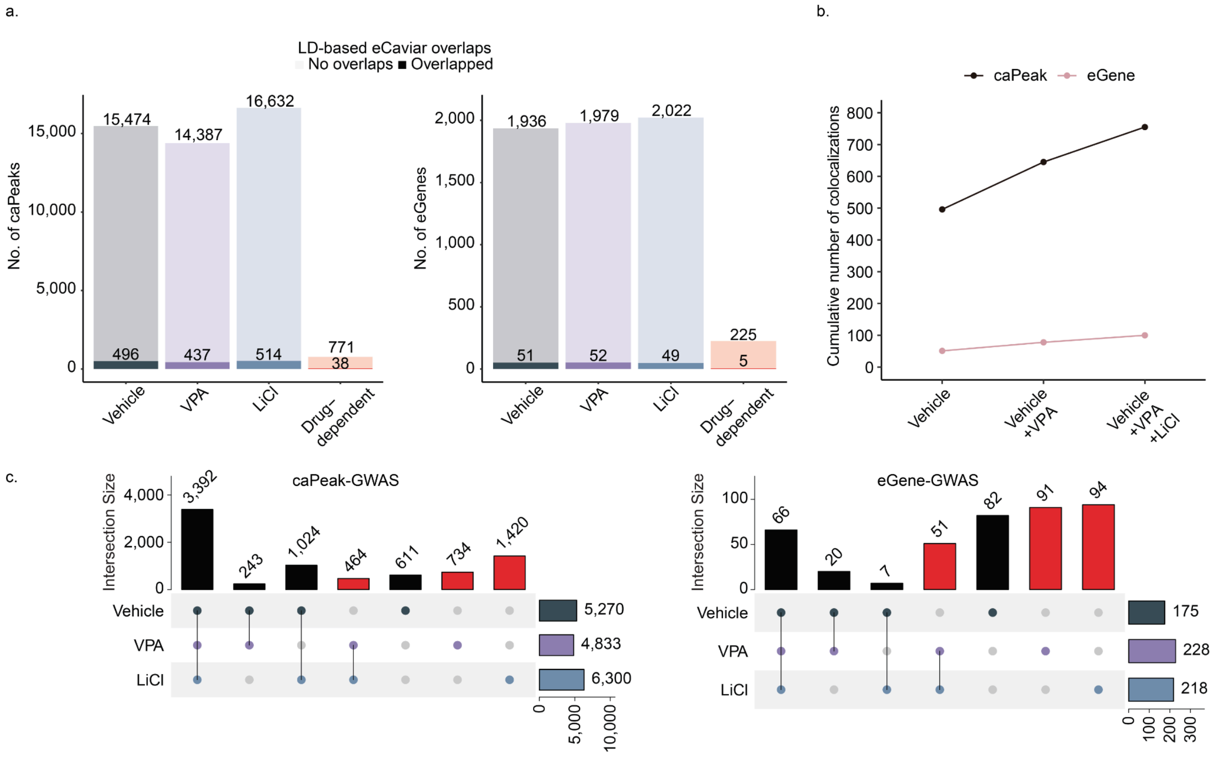

### Supplementary Fig. 8 | Colocalization of SUPT7L associated r-caQTL with average cortical thickness.

(a) Boxplot showing increased *SUPT7L* gene expression by rs6718978-G significant only after treatment by VPA. nvehicle = 76, nVPA = 78. (b) Boxplot showing a decrease in chromatin accessibility associated with rs6718978-G. An interaction effect between vehicle and VPA conditions significantly augments the effect. $n_{vehicle}$ = 76, $n_{VPA}$ = 75. P values for eQTLs were estimated by a linear mixed model using a two-sided test. caQTL P values were obtained using a likelihood-ratio test in RASQUAL (Methods). QTLs were adjusted for multiple testing correction by the Benjamini–Hochberg procedure. The lower and upper hinges in box plots correspond to the first and third quartiles, and center lines correspond to the median of the residuals. (c) Regional association plot depicting co-localization of average cortical thickness (top panel) GWAS with a VPA-responsive caQTL overlapping an eQTL modulating *SUPT7L* only after VPA treatment. From top to bottom: genomic coordinates and gene models, P values for brain-related GWAS, P values for QTLs discovered in this study and differentially accessible regions (DARs) within the locus.

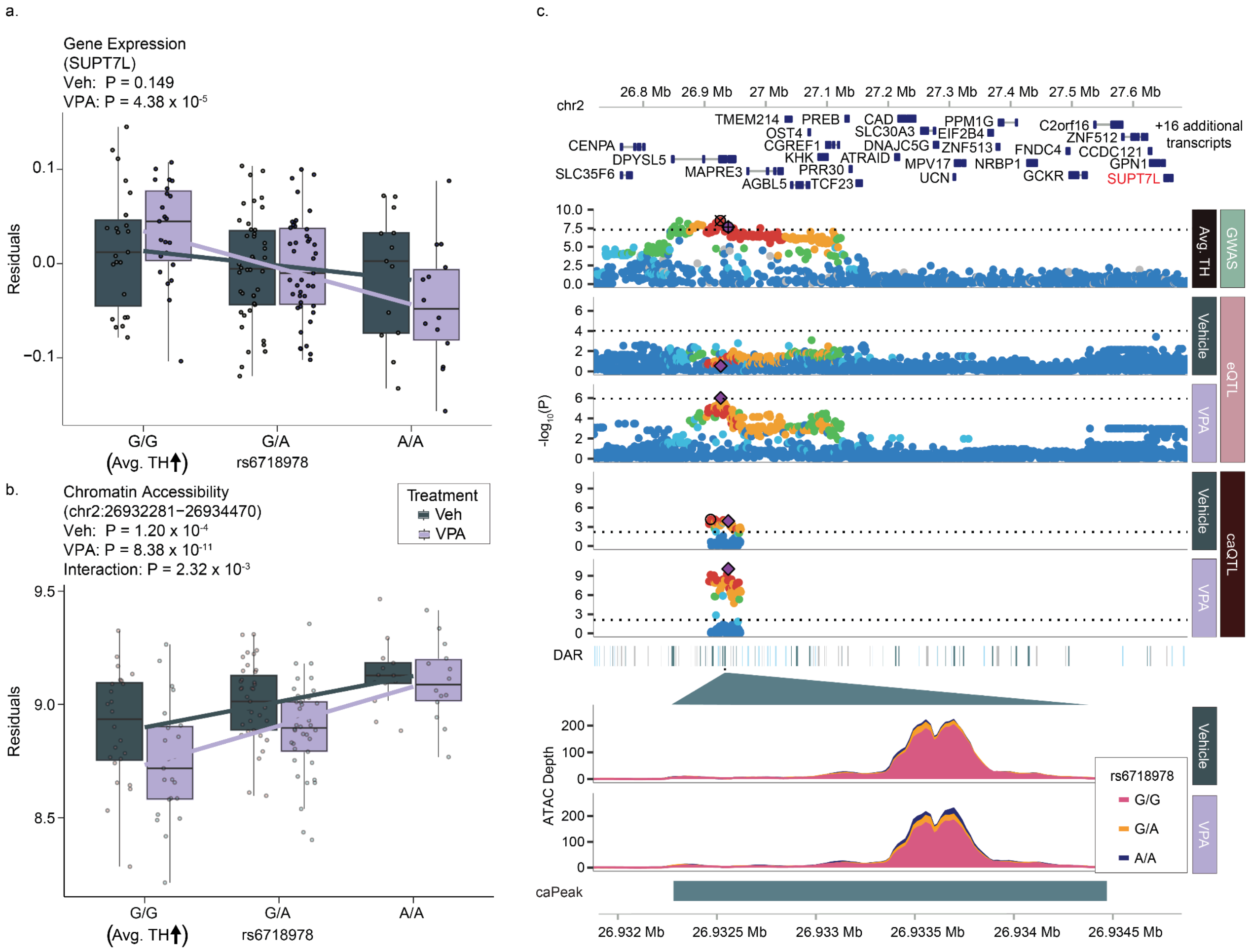

#### Supplementary Figure 9 | TWAS implicates condition-dependent gene-trait associations.

(a, b) TWAS to predict genes associated with ASD, Epilepsy, cortical surface area (SA), and thickness (TH)  (left) and bipolar (right). Each dot indicates the TWAS gene Z score in the context of VPA exposure (y-axis) or vehicle exposure (x-axis). Genes without significant heritability were set to TWAS Z-scores of zero. (c) Top 10 Genes identified by TWAS across GWAS phenotypes-condition (left: VPA and Vehicle; right: Li and Vehicle). Upward or downward triangles indicate positive or negative correlation to the GWAS phenotype, accordingly. (d) Pathways significantly enriched by genes associated with ASD, SA and TH after exposure to VPA. Numbers in parentheses indicate the intersection size, i.e., the number of DEGs overlapping with each pathway. K = KEGG, R = Reactome.

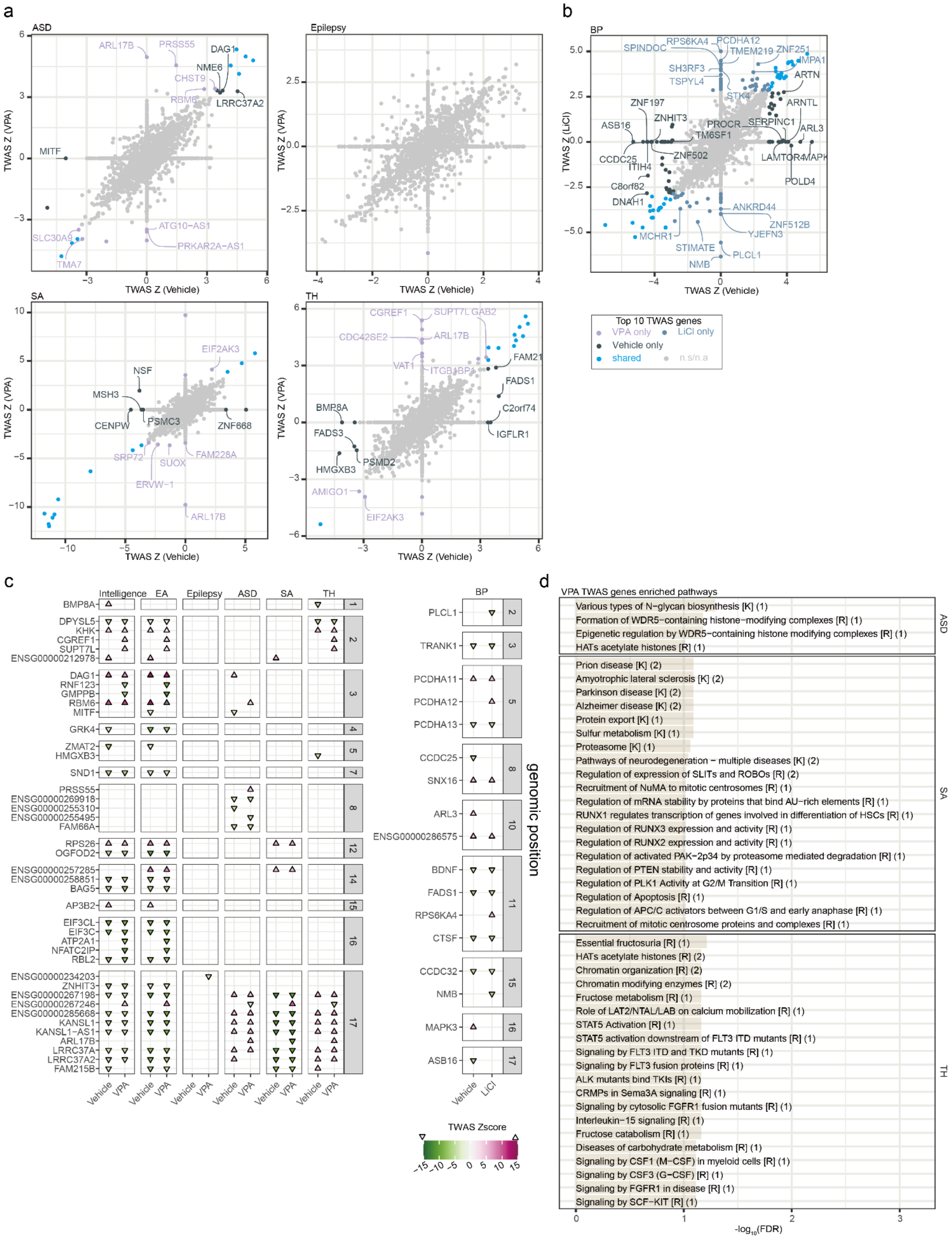
a
ASD
Epilepsy
SA
TH
TWAS Z (VPA)
TWAS Z (Vehicle)
ARL17B
PRSS55
DAG1
NME6
CHST9
RBM6
LRRC37A2
MITF
ATG10-AS1
PRKAR2A-AS1
SLC30A9
TMA7
EIF2AK3
NSF
MSH3
CENPW
PSMC3
ZNF668
SRP72
SUOX
FAM228A
ERVW-1
CGREF1
SUPT7L
GAB2
CDC42SE2
VAT1
ITGB3BP
FAM21
FADS1
C2orf74
IGFLR1
BMP8A
FADS3
PSMD2
HMGXB3
AMIGO1
b
BP
TWAS Z (LiCl)
SPINDOC
RPS6KA4
PCDHA12
TMEM219
ZNF251
IMPA1
SH3RF3
TSPYL4
ARTN
STK4
ZNF197
ZNHIT3
ARNTL
ASB16
PROCR
SERPINC1
TM6SF1
ARL3
CCDC25
ZNF502
LAMTOR4
MAPK
ITIH4
C8orf82
POLD4
DNAH1
ANKRD44
ZNF512B
MCHR1
YJEFN3
STIMATE
PLCL1
NMB
Top 10 TWAS genes
VPA only
LiCl only
Vehicle only
shared
n.s/n.a
c
Intelligence
EA
Epilepsy
ASD
SA
TH
BP
Vehicle
VPA
LiCl
genomic position
TWAS Zscore
d
VPA TWAS genes enriched pathways
ASD
Various types of N−glycan biosynthesis [K] (1)
Formation of WDR5−containing histone−modifying complexes [R] (1)
Epigenetic regulation by WDR5−containing histone modifying complexes [R] (1)
HATs acetylate histones [R] (1)
SA
Prion disease [K] (2)
Amyotrophic lateral sclerosis [K] (2)
Parkinson disease [K] (2)
Alzheimer disease [K] (2)
Protein export [K] (1)
Sulfur metabolism [K] (1)
Proteasome [K] (1)
Pathways of neurodegeneration − multiple diseases [K] (2)
Regulation of expression of SLITs and ROBOs [R] (2)
Recruitment of NuMA to mitotic centrosomes [R] (1)
Regulation of mRNA stability by proteins that bind AU−rich elements [R] (1)
RUNX1 regulates transcription of genes involved in differentiation of HSCs [R] (1)
Regulation of RUNX3 expression and activity [R] (1)
Regulation of RUNX2 expression and activity [R] (1)
Regulation of activated PAK−2p34 by proteasome mediated degradation [R] (1)
Regulation of PTEN stability and activity [R] (1)
Regulation of PLK1 Activity at G2/M Transition [R] (1)
Regulation of Apoptosis [R] (1)
Regulation of APC/C activators between G1/S and early anaphase [R] (1)
Recruitment of mitotic centrosome proteins and complexes [R] (1)
TH
Essential fructosuria [R] (1)
HATs acetylate histones [R] (2)
Chromatin organization [R] (2)
Chromatin modifying enzymes [R] (2)
Fructose metabolism [R] (1)
Role of LAT2/NTAL/LAB on calcium mobilization [R] (1)
STAT5 Activation [R] (1)
STAT5 activation downstream of FLT3 ITD mutants [R] (1)
Signaling by FLT3 ITD and TKD mutants [R] (1)
Signaling by FLT3 fusion proteins [R] (1)
ALK mutants bind TKIs [R] (1)
CRMPs in Sema3A signaling [R] (1)
Signaling by cytosolic FGFR1 fusion mutants [R] (1)
Interleukin−15 signaling [R] (1)
Fructose catabolism [R] (1)
Diseases of carbohydrate metabolism [R] (1)
Signaling by CSF1 (M−CSF) in myeloid cells [R] (1)
Signaling by CSF3 (G−CSF) [R] (1)
Signaling by FGFR1 in disease [R] (1)
Signaling by SCF−KIT [R] (1)
-log10(FDR)

Supplementary Tables
Supplementary Table 1: DARs
Supplementary Table 2: chromHMM Enrichment values
Supplementary Table 3: chromHMM Fisher values
Supplementary Table 4: TF enrichments
Supplementary Table 5: Pathway enrichments
Supplementary Table 6: LDSC
Supplementary Table 7: DEGs
Supplementary Table 8: Peak-gene correlation
Supplementary Table 9: caQTLs
Supplementary Table 10: eQTLs
Supplementary Table 11: r-caQTLs
Supplementary Table 12: r-eQTLs
Supplementary Table 13: caQTL-eQTL overlaps
Supplementary Table 14: r-QTL context discovered
Supplementary Table 15: eQTL context discovered and pLOEUF score
Supplementary Table 16: GWAS list
Supplementary Table 17: caQTL GWAS overlaps
Supplementary Table 18: eQTL GWAS overlaps
Supplementary Table 19: caQTL eCAVIAR colocalizations
Supplementary Table 20: eQTL eCAVIAR colocalizations
Supplementary Table 21: TWAS results
